# A Longitudinal Clinical Foundation Model on Nationwide Veteran Health Trajectories

**DOI:** 10.64898/2026.05.13.26353133

**Authors:** Rafael Zamora-Resendiz, Junqi Yin, Nathan A. Kimbrel, Jean C. Beckham, Million Veteran Program, Silvia Crivelli

## Abstract

We present VA-LLM, a 1.62-billion-parameter autoregressive transformer pre-trained from scratch on 1.74 trillion tokens of clinical text spanning 22 years of care for 13.8 million patients in the Veterans Health Administration, with mortality outcomes confirmed through the National Death Index for 7.8 million patients. In a retrospective–prospective evaluation on 107,555 withheld patients, VA-LLM achieved higher 5-year AUPRC than Llama-2 (7 billion parameters), BioGPT_large (1.57 billion parameters), and GatorTron (3.91 billion parameters), matching GatorTron’s 100,000-patient performance with only 10,000 labeled patients. In a clinical validation against the VA’s operational Care Assessment Need (CAN) score on 5.5 million patients one year beyond the pre-training corpus, VA-LLM achieved a 90-day mortality AUROC of 90.00% versus 87.74% (*p* < 0.001) and a 45% relative improvement in AUPRC; post-hoc recalibration recovered calibration comparable to CAN (Brier 0.0091 versus 0.0093) without sacrificing discrimination. Across 21 pre-training checkpoints, discriminative performance correlated more strongly with cumulative mortality experience (CME), the total person-years contributed by patients with confirmed deaths, than with token count (Δ*R*^2^ = 0.15; Williams *p* < 10^−6^). Performance plateaued once marginal cohorts added fewer confirmed deaths, even as pre-training loss continued to decrease. These findings suggest that the clinical composition of pre-training data, particularly the completeness of documented patient trajectories, correlates with predictive performance more closely than corpus size alone.

## Main

Large language models (LLMs) learn transferable representations from large text corpora^1^. Performance scales predictably with model and dataset size^2^, a relationship that has spurred substantial investment in training infrastructure and increasingly capable frontier models^3^. Yet the relationship between scale and clinical utility remains unclear. Clinicians managing large patient populations need risk indices to prioritize high-risk patients for outreach, goals-of-care discussions, and intensive case management^4^, yet labeled examples for many endpoints remain scarce even at national scale^5^. For outcomes such as suicide or rare diseases, a health system may have only thousands of confirmed events despite millions of patients with years of clinical documentation. Whether a foundation model pre-trained on clinical histories can enable label-efficient risk prediction depends not only on how much data the model has seen, but what clinical observations the data contain.

Patient trajectories are documented in clinical narratives. While structured electronic health records (EHR) fields capture billable events, discrete measurements, and coded diagnoses, clinical notes document dimensions of care that are under- represented in clinical codes^6^: how a patient presents, whether they are engaging with treatment, the clinician’s evolving assessment, and the reasoning behind changes in management. A single patient’s notes may document the initial presentation and comorbidity context, interval changes in functional status, shifts in clinician language from active treatment to prognostic hedging, and eventual transition to comfort-focused care. At the population level, these narratives accumulate into longitudinal records that trace individual trajectories through changing health states, often spanning years of care and, for patients who die, documenting the progression toward death. This trajectory information is precisely what outcome prediction requires, yet it is largely inaccessible to language models pre-trained on biomedical literature or general-domain text^7,8^.

The Department of Veterans Affairs (VA) operates the largest integrated healthcare system in the United States, with over 170 medical centers and 1,100 outpatient sites spanning every state^9^. The Veterans Health Administration’s (VHA) centralized EHR indexes more than 25 million patients. For a substantial subset, the record spans over a decade of continuous care across primary, specialty, surgical, psychiatric, and long-term settings, with clinical narratives linked to confirmed mortality outcomes.

The result is a uniquely dense record of disease progression at national scale. Prior clinical language modeling efforts have drawn on narrower resources: Epic’s Curiosity models train on structured events from 118 million patients but do not ingest clinical narrative^10^; NYUTron was pre-trained on notes from a single institution with 336,000 patients^11^; EHRSHOT evaluated structured-data representations on 6,739 patients^12^; and the largest published EHR scaling-law study trained on MIMIC-IV, a freely available intensive-care dataset of roughly 300,000 admissions, where model performance saturated at 28 million parameters due to data scarcity^13,14^. None combines population-scale patient volume, decade-spanning per-patient follow-up in clinical text, and confirmed decedent outcomes.

We hypothesized that language models pre-trained on longitudinal patient records learn representations of health trajectories that transfer to specific clinical endpoints through fine-tuning on small labeled cohorts, outperforming models pre-trained on general-domain text, biomedical literature, or clinical datasets with shorter per-patient follow-up^15^. We evaluated this hypothesis on all-cause mortality, which offers both clinical importance and outcome ascertainment independent of the documenting institution: mortality linkage with the Centers for Disease Control and Prevention’s National Death Index (NDI) provided confirmed outcomes for 7.8 million patients including time and cause of death. All-cause mortality integrates over many distinct disease processes, making it a natural summary measure of overall health decline and a practical starting point: if trajectory depth enables label-efficient discrimination for this broad outcome, the same approach can be adapted to scarcer, cause-specific deaths.

This work makes three contributions. First, we pre-trained VA-LLM, a 1.62-billion-parameter clinical language model, on 1.74 trillion tokens of VA clinical text from 13.8 million patients spanning 22 years. Second, we evaluated VA-LLM alongside three comparison foundation models in a retrospective–prospective framework, monitoring performance along pre-training checkpoints to investigate which properties of the training data, including observed patients, longitudinal coverage, and outcome resolution, correlate with predictive improvement. Third, we validated VA-LLM against the VA’s operational Care Assessment Need (CAN) score on 5.5 million patients, showing that text-derived representations match or exceed curated structured variables without feature engineering.

### Clinical Foundation Models

LLMs have shown broad utility in medical applications, including clinical concept retrieval, named entity recognition, and medical question answering^16^. Domain-specific models such as BioBERT^17^, ClinicalBERT^18^, BioMegatron^19^, BioGPT^7^, and GatorTron^8^ have shown that pre-training on biomedical literature and clinical text improves performance on natural language processing tasks. More recently, general-purpose LLMs have shown strong medical knowledge recall^20^, but their rapid scaling has outpaced the availability of sufficiently large healthcare corpora, motivating adaptation strategies such as continual pre-training^21^, fine-tuning^22^, and instruction tuning^23^.

A clinical foundation model extends this paradigm: a single pre-trained model whose representations transfer to diverse patient-care tasks^24^. Yet evaluations have focused almost exclusively on factual-recall benchmarks such as MedQA^25^, MedMCQA^26^, and PubMedQA^27^; these do not test whether a model can predict patient outcomes from longitudinal clinical records, a task that requires sensitivity to temporal disease progression rather than static medical knowledge.

Three gaps limit progress. First, existing benchmarks do not enforce temporal separation between training and evaluation^12^; even large-scale efforts such as Curiosity lack temporal holdout designs^10^, making it difficult to distinguish retrospective fit from prospective generalization. Second, how pre-training corpus characteristics shape predictive performance remains largely unexamined; recent scaling studies of structured EHR models report standard power-law relationships with parameters and tokens^10,13^ without identifying which clinical attributes of the training population drive improvement. Third, no clinical language model has been validated against an operational risk score at the population scale where such scores are actually used.

## Results

All evaluations follow a retrospective–prospective design^28^ (Supplementary Figure S3) in which models were fine-tuned exclusively on outcomes preceding a fixed calendar cutoff and evaluated on outcomes occurring after it, *so that no future information leaks into training*. We use two cutoffs to test progressively harder generalization. In the model comparison experiments (cutoff: January 1, 2017), fine-tuning and evaluation cohorts were drawn from patients excluded from pre-training; this setting tests whether pre-trained representations support adaptation to novel patients and prospective mortality discrimination on an independent held-out sample. In the clinical validation against the CAN score (cutoff: January 1, 2023), the evaluation window begins one year after the pre-training corpus ends; this setting tests whether learned representations further generalize to clinical contexts, practice patterns, and patient outcomes never observed during either pre-training or fine-tuning.

All models were adapted for binary mortality classification using Low-Rank Adaptation (LoRA)^29^ with approximately equivalent trainable parameters (∼100 million) to isolate the effect of pre-training data from architectural differences as closely as the design permits. Training configurations, available context window per patient, LoRA hyperparameters, and optimization settings were held constant across all models within each experiment, so that performance differences reflect differences in pre-training data rather than model architecture or training procedure (Table 2, Supplementary Table S2; see Methods).

### Pre-training on VA Clinical Text

VA-LLM was pre-trained on 1.74 trillion tokens derived from 6.31 terabytes of VA clinical text spanning January 2000 through December 2021. We ordered all nationally indexed patients by year of birth and partitioned them into 21 sequential cohorts, each containing approximately 600,000 patients with documented clinical text. Birth year serves as a label-free proxy for outcome resolution: older patients contribute outcome-resolved trajectories through confirmed death, whereas younger, actively followed patients contribute right-censored records with unknown vital status. No mortality labels were used during pre-training. This ordering causes corpus-level quantities that normally co-vary, including tokens, patients, person-years, confirmed deaths, and cumulative mortality experience (CME; the total person-years contributed by patients with confirmed death), to decouple across checkpoints, isolating variation in outcome resolution from simple increases in scale. Its complement, cumulative survival experience (CSE), captures person-years from patients without confirmed death. CME and CSE are measured in the same units from the same corpus; they differ only in whether the trajectory is outcome-resolved. If corpus volume alone governs performance, both should predict equally well.

Early cohorts (c1–c5, World War II–era Veterans) have mortality rates exceeding 90% and contribute a high density of outcome-resolved trajectories; later cohorts (c16–c21, Gulf War– and Afghanistan–era Veterans) have mortality rates below 35% and contribute proportionally more right-censored courses (Supplementary Table S1). Each pre-training checkpoint corresponds to a realistic deployed model state after *k* incremental updates on successive one-million–Veteran enrollment cohorts, giving the 21-checkpoint series direct operational rather than purely methodological meaning.

Under the null hypothesis that performance scales with token volume alone, both pre-training loss and downstream discrimination should improve primarily as a function of token count. Pre-training loss decreased across all checkpoints, consistent with this null. However, downstream discriminative performance plateaued after checkpoint 16, inconsistent with the token-volume-only null for downstream discrimination. Because the birth-year curriculum causes candidate variables, including patients, confirmed deaths, patient-years, and cumulative mortality experience, to grow at different rates than tokens (Supplementary Table S1), their relative contributions can be distinguished statistically (Supplementary Note S3.4).

To assess generalization to unseen patients, we withheld 279,125 patients from pre-training as a 2% stratified sample of the full VA population. The sample was matched on age, sex, race, ethnicity, mortality status, and total encounters to preserve demographic and healthcare utilization representativeness (Table 1). This held-out cohort reflects the historical VA patient population from 2000–2021: predominantly male (89.1%), White (55.3%), and non-Hispanic (68.3%), with a median of 52 encounters per patient and an overall mortality rate of 55.5% through December 2021. All fine-tuning and model selection were conducted exclusively in this held-out cohort.

**Table 1.**
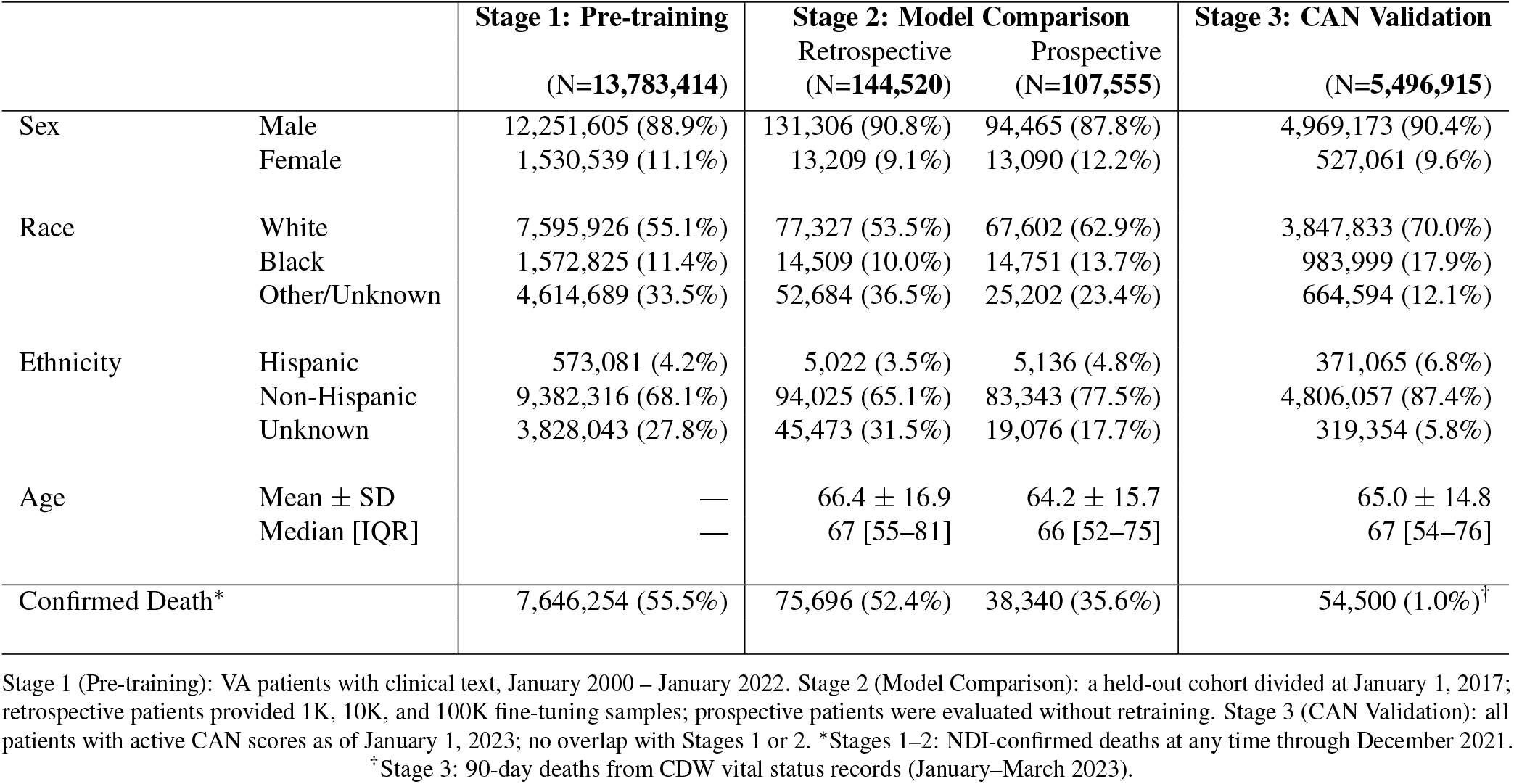
Cohort characteristics across study phases. The study comprises three stages: (1) pre-training on the full VA clinical corpus; (2) model comparison, in which a held-out cohort is split into a retrospective fine-tuning pool and a prospective evaluation set; and (3) CAN validation against the VA’s operational mortality score. Data from the VA’s TIU database linked to CDC’s NDI.

### All-Cause Mortality Prediction

Within the held-out cohort, patients with any clinical text before January 1, 2017 were eligible for the model comparison experiments. Among these, patients with outcomes on or before that date formed the retrospective fine-tuning set (*n* = 144,520), and patients with outcomes after that date formed the prospective evaluation cohort (*n* = 107,555). Outcome dates for deceased patients corresponded to NDI-confirmed death dates. Living controls were birth-date matched to deceased cases and assigned a prediction time point at the same age as their matched case’s death, ensuring that cases and controls had comparable lengths of clinical history available for prediction. Class imbalance varied by prediction horizon, with 1.36% of the prospective evaluation cohort deceased at 90 days, 2.64% at 180 days, 5.26% at 1 year, and 35.67% at 5 years.

Table 2 presents prospective performance across VA-LLM checkpoints and comparison models at four horizons (90-day, 180-day, 1-year, 5-year). All models were fine-tuned on a single binary label (alive or dead at one week before the outcome date) using case-control sets of 1K, 10K, and 100K patients. This design tests how little labeled data is needed to produce a useful risk discriminator from pre-trained representations, the label-efficient adaptation scenario motivating this work. Five-year Area Under the Precision-Recall Curve (AUPRC) serves as the primary comparison metric because the five-year horizon yields the highest event rate among the four windows, providing the most statistical power to distinguish models on the precision-recall trade-off. Area Under the Receiver Operating Characteristic curve (AUROC), which is more stable at low event rates, is reported across all four horizons to show how discrimination changes as the prediction window widens.

**Table 2.**
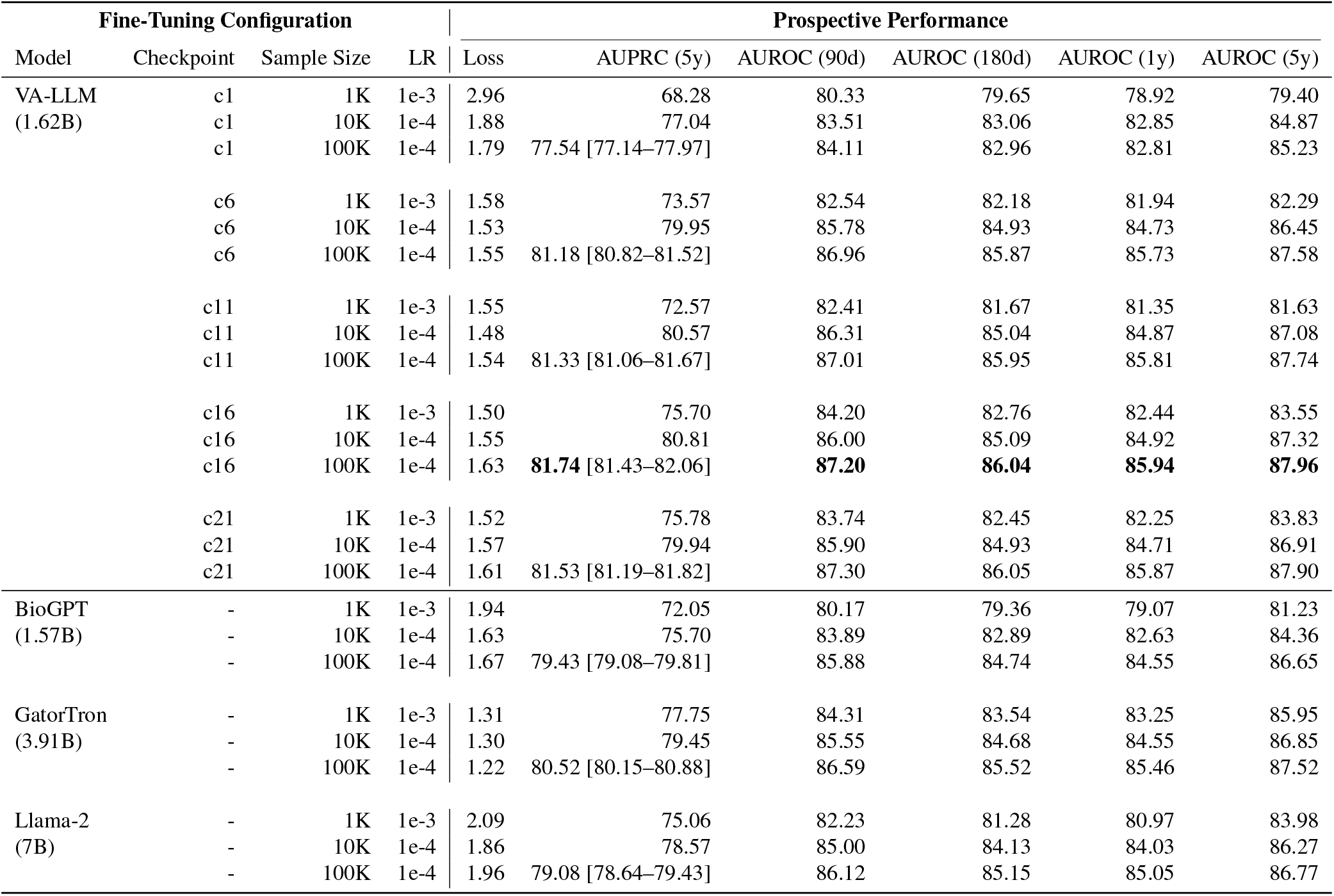
Prospective all-cause mortality prediction performance. AUROC and AUPRC at 90-day, 180-day, 1-year, and 5-year horizons from January 1, 2017 on 107,555 withheld patients. VA-LLM checkpoints (c1–c21) are evaluated alongside BioGPT_large, GatorTron_medium, and Llama-2 at 1K, 10K, and 100K fine-tuning sample sizes. AUPRC is the primary metric given class imbalance. Bootstrap 95% CIs (1,000 resamples, percentile method) are reported for 100K rows.

#### VA-LLM outperforms larger models pre-trained on broader corpora

We benchmarked VA-LLM against three foundation models chosen to represent the major sources of pre-training data currently being adapted to clinical applications: general-domain text (Llama-2, 7 billion parameters), biomedical literature (BioGPT_large, 1.57 billion parameters), and clinical notes from a single health system (GatorTron_medium, 3.91 billion parameters; University of Florida). These were the largest openly available models in each category that could be deployed within the VA’s secure computing environment when the evaluation pipeline was frozen (July 2024); more recent architectures were not yet deployable on the constrained infrastructure (see Methods). The experiment is designed to isolate the effect of pre-training corpus provenance under matched adaptation conditions, not to benchmark against the latest model families. All models were fine-tuned with equivalent LoRA configurations on the same held-out cohort, so that performance differences reflect pre-training data rather than architectural or optimization choices.

At 100,000 fine-tuning samples, VA-LLM (checkpoint 16) achieved the highest 5-year AUPRC (81.74% [95% CI: 81.43–82.06]), followed by GatorTron (80.52% [80.15–80.88]), BioGPT (79.43% [79.08–79.81]), and Llama-2 (79.08% [78.64–79.43]). Model rankings shifted with sample size. With 1,000 fine-tuning patients, GatorTron (77.75%) outperformed VA-LLM (75.70%). This advantage reversed at 10,000 patients, where VA-LLM (80.81%) exceeded GatorTron (79.45%), and held at 100,000. BioGPT achieved the lowest AUPRC across all sample sizes (72.05% to 79.43%). VA-LLM also required 2.5-fold less fine-tuning compute than GatorTron (15.9 vs. 39.0 hours on 100,000 patients; Supplementary Table S2). At 10,000 fine-tuning patients, VA-LLM (80.81%) already exceeded GatorTron’s 100,000-patient performance (80.52%), achieving comparable discrimination at a fraction of the labeled data and compute (Figure 2a).

**Figure 1.**
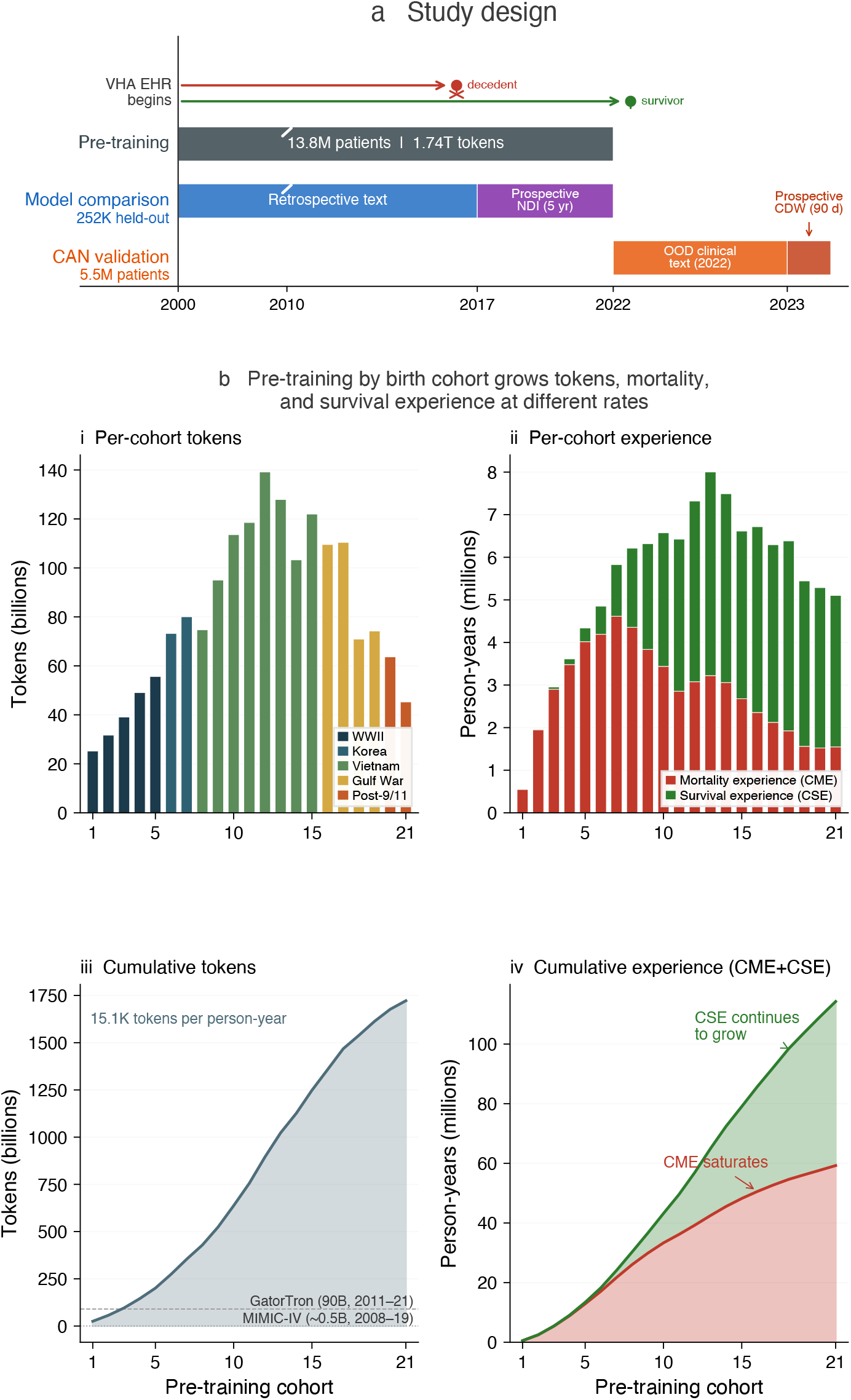
Study design and pre-training data. (a) Three-stage study timeline. Stage 1: pre-training on 13.8 million VHA patients (2000–2022) organized into 21 birth-year cohorts. Stage 2: fine-tuning with NDI-linked five-year mortality labels on 252,000 held-out patients. Stage 3: prospective-style validation against the CAN score on 5.5 million patients seen in January 2023. (b) Pre-training corpus properties by birth cohort. Per-cohort token volume (i) and person-years decomposed into cumulative mortality experience (CME) and cumulative survival experience (CSE) (ii); cumulative tokens (iii) and cumulative experience (iv). Token volume grows, but CME saturates while CSE continues to grow, reflecting the declining mortality rate in younger cohorts.

**Figure 2.**
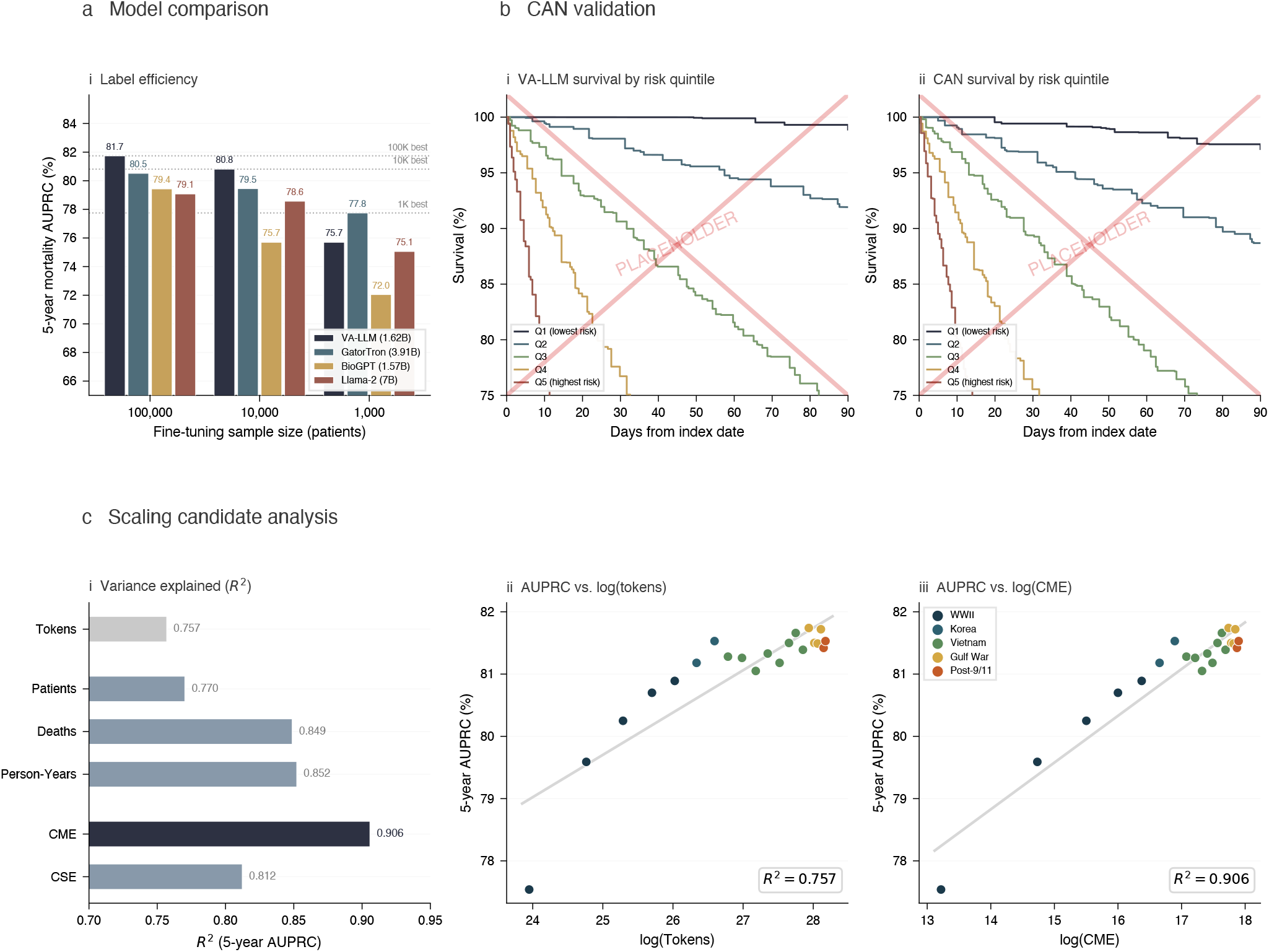
Performance and scaling analysis. (a) Label efficiency: five-year AUPRC for VA-LLM, GatorTron, BioGPT, and Llama-2 fine-tuned on 100,000, 10,000, and 1,000 patients. VA-LLM matches or exceeds all comparators at 100,000 and 10,000 patients. (b) Prospective CAN validation: 90-day Kaplan–Meier survival curves for 5.5 million patients stratified by VA-LLM risk quintile (i) and CAN score quintile (ii). (c) Scaling candidate analysis: *R*^2^ of log-linear fits between six candidate scaling variables and five-year AUPRC across 21 pre-training checkpoints (i); AUPRC versus log-transformed token count (*R*^2^ = 0.757) (ii) and CME (*R*^2^ = 0.906) (iii). Points are colored by military-service era. In this curriculum, CME explains substantially more variance than any other candidate.

#### Pre-training narrows the gap between small and large labeled cohorts

The AUPRC gap between 1K and 100K fine-tuning narrowed from 9.3 percentage points at checkpoint 1 (68.28% vs. 77.54%) to 6.0 points at checkpoint 16 (75.70% vs. 81.74%) and 5.8 points at checkpoint 21 (75.78% vs. 81.53%). With only 1K patients, AUPRC rose from 68.28% (checkpoint 1) to 75.78% (checkpoint 21), approaching the 100K baseline at checkpoint 1 (77.54%).

#### Discrimination improves up to ten million patients, then plateaus

Five-year AUPRC rose 4.2 percentage points as pre-training exposure increased from 543,000 to 9.95 million patients, then plateaued. At checkpoint 1 (543K patients, 25B tokens), VA-LLM achieved 77.54% AUPRC with 100K fine-tuning samples; by checkpoint 16 (9.95M patients, 1.36T tokens), performance reached 81.74%. Beyond checkpoint 16, further exposure yielded no additional gain: checkpoint 21 (13.1M patients, 1.72T tokens) achieved 81.53%. This saturation aligns with the trajectory gradient described above: the cohorts added after checkpoint 16 contribute proportionally fewer outcome-resolved trajectories, consistent with the hypothesis that outcome resolution rather than raw corpus volume is associated with improvement. The same pattern held across horizons: 90-day AUROC improved from 84.11% (checkpoint 1) to 87.30% (checkpoint 21), and 1-year AUROC from 82.81% to 85.87%.

#### Cumulative mortality experience is the strongest correlate of discrimination

To test whether clinical composition, rather than corpus size alone, predicts discriminative performance, we computed six candidate scaling variables for each of the 21 checkpoints: total tokens, total patients, total person-years, number of confirmed deaths, CME, and CSE (Figure 2c; see Methods).

At 100,000 fine-tuning patients, five-year AUPRC increased with all six variables, but the strength of association differed. Total tokens explained 76% of the variance in AUPRC across checkpoints (*R*^2^ = 0.76), total patients 77% (*R*^2^ = 0.77), person-years 85% (*R*^2^ = 0.85), number of deaths 85% (*R*^2^ = 0.85), CSE 81% (*R*^2^ = 0.81), and CME 91% (*R*^2^ = 0.91). CSE explained less variance than CME despite capturing more total person-years in later checkpoints, consistent with outcome resolution rather than follow-up volume being the more informative correlate in this curriculum.

Because all six variables grow together as cohorts are added, simple *R*^2^ comparisons cannot establish whether the differences are statistically meaningful. We applied the Williams test for comparing correlated predictors on the same observations. CME was a significantly stronger correlate of AUPRC than token count (*r*_CME_ = 0.95 versus *r*_tokens_ = 0.87; Williams *t* = 8.18, *d f* = 18, *p <* 10^*Δ*6^), explaining 15 additional percentage points of variance (Δ*R*^2^ = 0.15) despite the two variables sharing 97% of their variation (*r* = 0.97). Because all candidate variables increase together under the birth-year curriculum, these correlational results cannot isolate CME as a causal factor; we discuss the co-varying confounds and the ablations needed to resolve them below. A similar ordering was observed at 10,000 fine-tuning patients, though with fewer sampled checkpoints and correspondingly less statistical power (Supplementary Table S6).

### Clinical Validation Against CAN Score

The preceding analyses serve as model selection and scaling candidate analysis: they establish that pre-training on longitudinal trajectories with confirmed outcomes improves both discrimination and sample efficiency relative to broader but clinically shallow corpora, under matched conditions where all models share the same January 2017 cutoff. Because that cutoff falls within the pre-training period, the model may have encountered clinical text from the same era during pre-training, and could therefore exploit period-specific documentation conventions, formulary practices, or facility-level language patterns that correlate with outcomes in that window but do not generalize beyond it. The prospective test of whether the learned representations capture generalizable prognostic structure rather than such temporal artifacts requires evaluation on data entirely outside the pre-training window. We tested this by predicting 90-day mortality as of January 1, 2023, one year beyond the end of the pre-training corpus (January 2022), on patients whose clinical context used for the prediction was never seen during either pre-training or fine-tuning.

We benchmark against the Care Assessment Need (CAN) score, the VA’s national operational standard for mortality risk stratification^30^. CAN is a logistic regression model that derives calibrated 90-day mortality probabilities from curated structured EHR variables, including demographics, diagnoses, utilization patterns, laboratory values, and vital signs. Developed and iteratively refined over more than a decade of deployment, CAN represents the baseline discrimination the VA can achieve from structured data alone. It is routinely used by clinical teams across all VA facilities to prioritize care management, palliative care referrals, and resource allocation^4^, making it a rigorous clinical benchmark rather than a research baseline.

We generated VA-LLM predictions (checkpoint 16, 100K fine-tuning samples) for 5.5 million patients with CAN scores computed as of January 1, 2023 and any clinical text during 2022, none of whom were included in fine-tuning. To match CAN’s operational input window, VA-LLM used only clinical text from the preceding year (see Methods). This restriction ensures a fair comparison on identical temporal scope; whether incorporating earlier longitudinal history improves discrimination beyond this window is examined in the model comparison experiments. Table 3 summarizes discrimination, calibration, and risk stratification metrics for 90-day mortality.

**Table 3.** Clinical validation: VA-LLM (checkpoint 16, 100K fine-tuning) versus the VA’s operational CAN score for 90-day mortality on 5,496,915 patients evaluated from January 1, 2023. Bootstrap 95% CIs from 1,000 patient-level resamples (percentile method).

|  | VA-LLM | CAN Score |
| --- | --- | --- |
| *Discrimination* |  |  |
| AUROC (% , 95% CI) | 90.00 [89.90–90.11] | 87.74 [87.60–87.86] |
| AUPRC (% , 95% CI) | 14.45 [14.17–14.68] | 9.99 [9.79–10.19] |
| DeLong test | | $z = 41.70, p < 0.001$ |
| *Calibration (uncalibrated)* |  |  |
| Brier score | 0.1554 | 0.0093 |
| Calibration slope | 0.3490 | 0.9949 |
| *Calibration (isotonic, 5% fit)* |  |  |
| Brier score | 0.0091 | — |
| Calibration slope | 0.9725 | — |
| *Risk stratification* |  |  |
| HR, Q5 vs Q1 (95% CI) | 189.48 [167.16–214.77] | 124.13 [111.81–137.82] |
| Log-rank $p$ | < 0.001 | < 0.001 |

#### Discrimination

VA-LLM achieved an AUROC of 90.00% [95% CI: 89.90–90.11%] and AUPRC of 14.45% [95% CI: 14.17–14.68%]; CAN achieved an AUROC of 87.74% [95% CI: 87.60–87.86%] and AUPRC of 9.99% [95% CI: 9.79–10.19%] (DeLong *z* = 41.70, *p <* 0.001). The 45% relative improvement in AUPRC is notable because precision-recall performance is most informative at low event rates, where a model must maintain precision as it increases recall into a population dominated by survivors. VA-LLM achieves comparable or superior discrimination from a single classification head applied to frozen pre-trained representations of unstructured clinical text, requiring no variable selection, feature engineering, or domain-specific data pipelines.

#### Subgroup Discrimination

Table 4 reports VA-LLM and CAN AUROC stratified by sex, race, ethnicity, and age group on the CAN validation cohort, using the recalibrated VA-LLM scores described in Supplementary Note S3.1. VA-LLM maintained higher AUROC than CAN in every stratum, with no reversals or crossings of confidence intervals. The advantage was consistent across race (White 0.897 vs. 0.873; Black 0.902 vs. 0.881; Other/Unknown 0.906 vs. 0.888) and ethnicity (Hispanic 0.926 vs. 0.909; Not Hispanic 0.898 vs. 0.875), with no stratum showing a differential deficit. Both models showed declining AUROC with age, consistent with the greater clinical heterogeneity and higher baseline mortality in older cohorts; the VA-LLM advantage persisted in the ≥85 stratum (0.756 vs. 0.730) despite the universal degradation of discrimination at a 4.80% event rate. Full results including AUPRC and Brier score by stratum are reported in Supplementary Note S3.2.

**Table 4.**
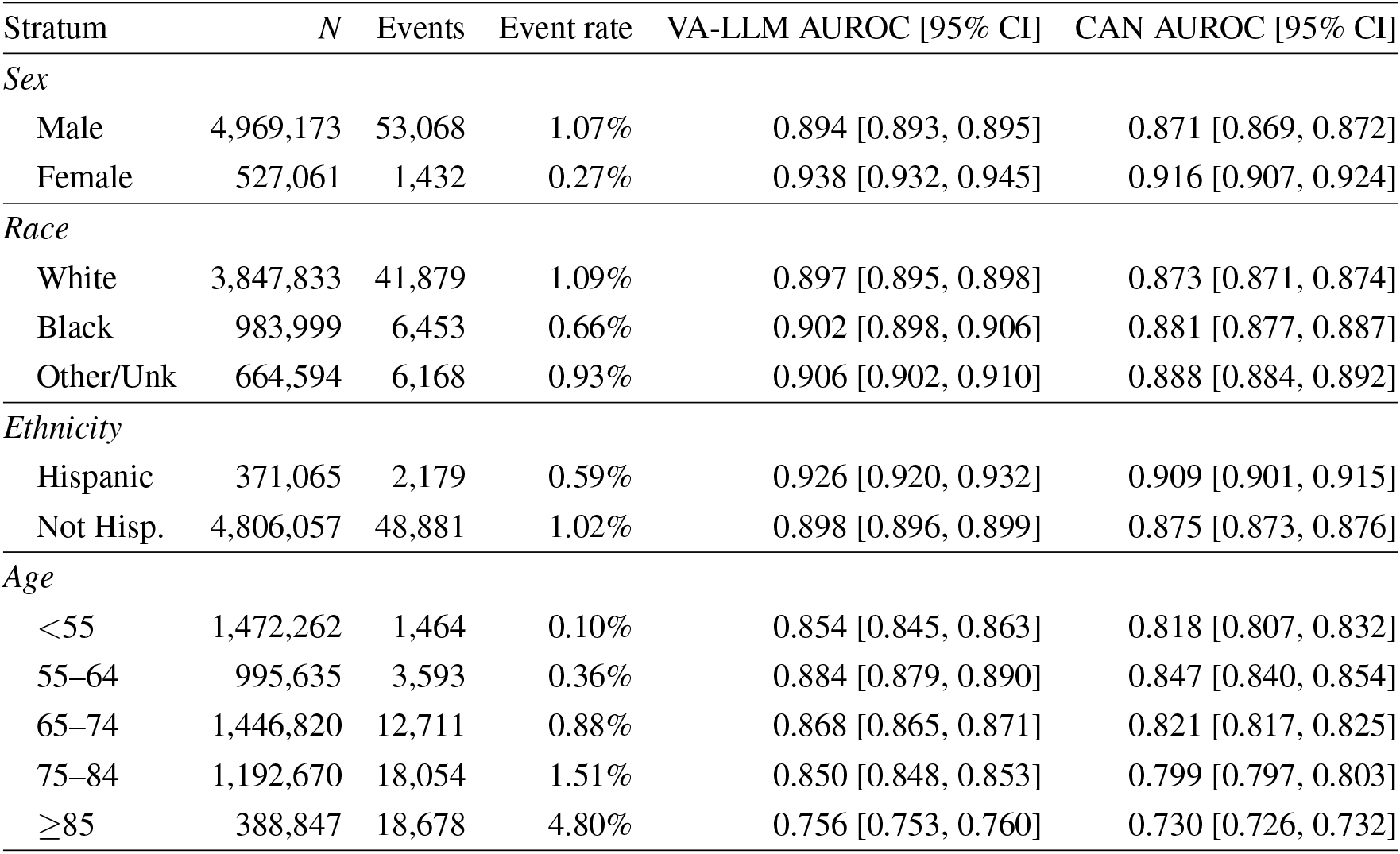
VA-LLM and CAN 90-day AUROC by demographic subgroup on the CAN validation cohort (January 2023). VA-LLM values use the recalibrated scores from Supplementary Note S3.1. Full results including AUPRC and Brier score are reported in Supplementary Table S4.

| Stratum | N | Events | Event rate | VA-LLM AUROC [95% CI] | CAN AUROC [95% CI] |
| --- | --- | --- | --- | --- | --- |
| *Sex* |  |  |  |  |  |
| Male | 4,969,173 | 53,068 | 1.07% | 0.894 [0.893, 0.895] | 0.871 [0.869, 0.872] |
| Female | 527,061 | 1,432 | 0.27% | 0.938 [0.932, 0.945] | 0.916 [0.907, 0.924] |
| *Race* |  |  |  |  |  |
| White | 3,847,833 | 41,879 | 1.09% | 0.897 [0.895, 0.898] | 0.873 [0.871, 0.874] |
| Black | 983,999 | 6,453 | 0.66% | 0.902 [0.898, 0.906] | 0.881 [0.877, 0.887] |
| Other/Unk | 664,594 | 6,168 | 0.93% | 0.906 [0.902, 0.910] | 0.888 [0.884, 0.892] |
| *Ethnicity* |  |  |  |  |  |
| Hispanic | 371,065 | 2,179 | 0.59% | 0.926 [0.920, 0.932] | 0.909 [0.901, 0.915] |
| Not Hisp. | 4,806,057 | 48,881 | 1.02% | 0.898 [0.896, 0.899] | 0.875 [0.873, 0.876] |
| *Age* |  |  |  |  |  |
| <55 | 1,472,262 | 1,464 | 0.10% | 0.854 [0.845, 0.863] | 0.818 [0.807, 0.832] |
| 55–64 | 995,635 | 3,593 | 0.36% | 0.884 [0.879, 0.890] | 0.847 [0.840, 0.854] |
| 65–74 | 1,446,820 | 12,711 | 0.88% | 0.868 [0.865, 0.871] | 0.821 [0.817, 0.825] |
| 75–84 | 1,192,670 | 18,054 | 1.51% | 0.850 [0.848, 0.853] | 0.799 [0.797, 0.803] |
| ≥85 | 388,847 | 18,678 | 4.80% | 0.756 [0.753, 0.760] | 0.730 [0.726, 0.732] |

#### Calibration

VA-LLM’s discrimination and calibration must be read as two distinct properties. *Uncalibrated*, VA-LLM produced strong risk rankings (AUROC 0.900, AUPRC 0.145) but poorly calibrated absolute probabilities: Brier score 0.1554, calibration slope 0.349, intercept Δ4.82, reflecting the expected behavior of a model fine-tuned on balanced case–control samples rather than observed population event rates. CAN, trained directly on population mortality rates, achieved near-perfect calibration (Brier 0.0093, slope 0.995, intercept Δ0.053). *After recalibration* with isotonic regression fit on 5% of the validation cohort, VA-LLM matched CAN’s calibration (Brier 0.0091, slope 0.973) while preserving its superior discrimination (AUROC 0.900, AUPRC 0.145; Table 3 and Supplementary Note S3.1). The calibration gap is therefore consistent with a training-objective mismatch rather than poor rank-order signal; standard post-hoc recalibration on a small operational sample is sufficient to recover it.

#### Risk Stratification

Kaplan–Meier survival curves based on VA-LLM risk quintiles showed clear monotonic separation across all five strata (Figure 2b; log-rank *p <* 0.001), with observed 90-day mortality rates increasing from 0.02% in the lowest-risk quintile (Q1; 246 deaths among 1,099,822 patients) to 4.15% in the highest-risk quintile (Q5; 45,617 deaths among 1,099,226 patients). CAN quintiles showed a similar monotonic gradient (Q1: 0.03%, Q5: 3.92%) but with less concentration of events at the extremes. VA-LLM assigned 83.7% of all 90-day deaths to Q5, compared with 79.1% for CAN, a difference of approximately 2,500 additional deaths correctly stratified into the highest-risk group across 5.5 million patients. The Q5-versus-Q1 hazard ratio was 189.48 for VA-LLM [95% CI: 167.16–214.77] and 124.13 for CAN [95% CI: 111.81–137.82], both significant at *p <* 0.001.

Reclassification analysis identified 187,624 patients whom CAN ranked as low risk (Q1–Q2) but VA-LLM ranked as high risk (Q4–Q5); 523 of these patients died within 90 days (observed mortality 0.28%). In the converse group, 153,043 patients whom CAN ranked as high risk but VA-LLM ranked as low risk, 205 died (observed mortality 0.13%). The two discordant groups differed sharply in age: patients reclassified upward by VA-LLM had a median age of 51.7 years, more than 22 years younger than those reclassified downward (73.9 years), consistent with VA-LLM and CAN capturing partially distinct prognostic information, particularly in younger Veterans where structured risk factors may be less informative (Supplementary Note S3.3). We hypothesize that documentation preceding the upward-reclassified deaths is enriched for textual signals under-represented in structured variables, such as functional decline, goals-of-care discussions, and patterns of treatment non-engagement. Model-guided interpretability analyses to test this hypothesis are the focus of future work.

## Discussion

The preceding results support a straightforward conclusion: what the pre-training corpus contains matters for clinical prediction at least as much as how large it is. VA-LLM discriminated prospective mortality risk one year beyond any data it had seen, matched a decade-old operational score on 5.5 million patients, and achieved comparable performance to a larger clinical model with ten-fold fewer labels, all from a corpus whose distinguishing feature is not volume but the proportion of patients whose complete course of illness, through confirmed death, is documented. GatorTron, the closest clinical-text comparator, was pre-trained on over 90 billion tokens from a single academic health system^8^; the label-efficiency advantage we observe suggests that longitudinal depth and outcome resolution compensate for a larger architecture. More broadly, no clinical language modeling effort, including Curiosity^10^, NYUTron^11^, and structured-event scaling studies on MIMIC-IV^13^, has drawn on a corpus of comparable longitudinal density, case-mix diversity, and outcome resolution.

### Clinical utility and deployment considerations

The reclassification results suggest that clinical notes contain prognostic information that CAN’s 36 curated structured variables^30^ capture only partially. This degree of risk separation could improve case-finding for intensive outreach, palliative care referral, and proactive goals-of-care discussions. For population-level triage, where the goal is to rank patients for clinical review, discrimination is the relevant criterion; standard post-hoc recalibration on a modest operational cohort would be required before outputs could serve as absolute risk estimates (Supplementary Note S3.1). Because VA-LLM and CAN draw on different data modalities, their errors may be partially independent, and combining both scores could further improve decision support. The model’s size (1.62 billion parameters) permits periodic batch scoring of millions of patients on standard GPU infrastructure, and the same backbone can be adapted to new endpoints as labels become available.

### Pre-training improves label efficiency

The ten-fold reduction in labeling burden demonstrated above matters because for many clinical outcomes, including suicide and rare-disease mortality, assembling 100,000 well-labeled patients is not realistic even at national scale.

The loss and AUPRC trajectories across pre-training checkpoints (Figure 3) reveal an abrupt transition in fine-tuning dynamics. In an early regime, additional pre-training reduced evaluation cross-entropy and improved AUPRC together; beyond a transition point, further pre-training raised evaluation cross-entropy while AUPRC plateaued or continued modest gains. One plausible explanation is that additional pre-training acts as an increasingly strong regularizer on the fine-tuning objective^31^: as the model accumulates more clinical priors, it resists conforming to the labeled sample’s empirical distribution and instead preserves the representations acquired during pre-training. Under this view, cross-entropy increases because predicted probabilities no longer track the case–control base rate of the fine-tuning set, while discrimination remains stable or improves because the rank ordering of patient risk is governed by pre-trained representations rather than fine-tuning statistics. Alternative explanations include distributional shifts across birth-year cohorts, label noise, and optimization dynamics; distinguishing among these mechanisms will require controlled experiments that vary pre-training data composition at fixed labeled cohorts. This regularization account predicts that smaller labeled cohorts, which are more susceptible to overfitting, should benefit from stronger pre-trained priors for longer before the regime transition occurs. The data confirm this: the transition shifted from checkpoint 7 with 100,000 fine-tuning patients to checkpoint 11 with 10,000 and checkpoint 16 with 1,000. At checkpoint 16, the 1,000-patient model achieved lower cross-entropy (1.50) than the 100,000-patient model (1.63) despite substantially lower AUPRC (75.70% vs. 81.74%), confirming that fine-tuning loss and discrimination are governed by different dynamics of the pre-trained backbone.

**Figure 3.**
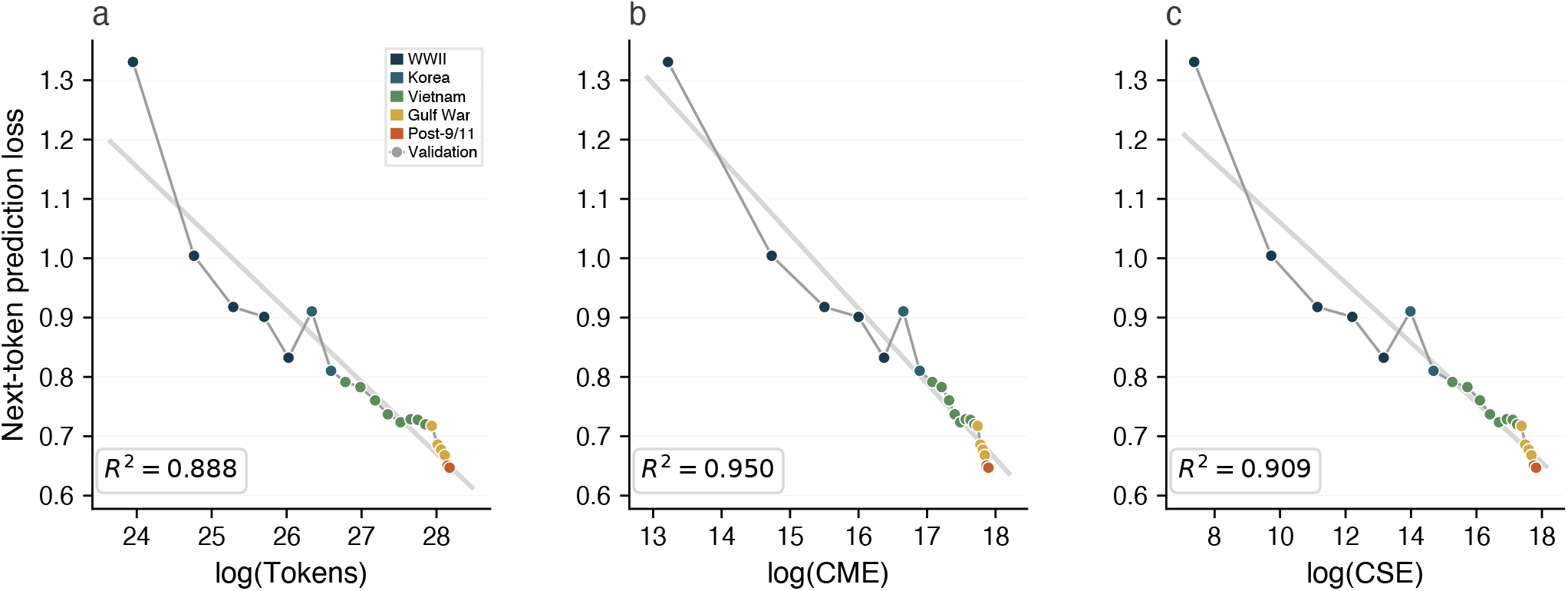
Pre-training validation loss across 21 checkpoints, indexed by three candidate scaling variables: cumulative tokens (a), CME (b), and CSE (c). Each panel shows era-colored markers with a linear fit and *R*^2^. All three variables predict validation loss, but the tightness of fit varies: CME and CSE yield higher *R*^2^ than token count, reflecting the non-uniform relationship between corpus volume and clinical content across the birth-year curriculum. The full scaling grid including prospective fine-tuning loss and AUPRC is shown in Supplementary Figure S4.

### Scaling clinical foundation models

Conventional neural scaling laws describe LLM performance as a power-law function of parameters, tokens, and compute^2,3^. Recent studies of structured EHR foundation models confirm that these relationships hold for clinical event sequences: Zhang et al. demonstrated log-linear scaling on MIMIC-IV patient timelines with performance saturating at 28 million parameters due to data scarcity^13^, while Epic’s Curiosity models scaled to one billion parameters and 151 billion tokens from 118 million patients^10^. Both operate on structured medical events and report scaling behavior consistent with standard token-parameter tradeoffs. Whether these tradeoffs predict performance on clinical prediction tasks, as opposed to pre-training loss, has not been tested.

Importantly, CME is a summary statistic of the pre-training corpus, not a filter on training data: all patients, including those with right-censored records, contribute clinical text during pre-training. CME quantifies how much documented clinical care preceding a confirmed death the model has encountered.

Three features of the scaling pattern are difficult to reconcile with raw corpus size as the operative variable. First, later checkpoints are temporally closer to the 2017 evaluation cutoff than earlier checkpoints, yet AUPRC does not track this closeness, arguing against recency of documentation as the primary driver. Second, pre-training cross-entropy continued to decrease from checkpoint 16 through checkpoint 21 while AUPRC held flat, indicating that additional tokens produced linguistic improvement without prognostic gain. Third, the cohorts added beyond checkpoint 16 contribute proportionally fewer confirmed deaths, so that marginal AUPRC tracks marginal CME rather than marginal tokens.

These patterns are nonetheless inherently correlational. The birth-year curriculum jointly varies patient volume, age distribution, service era, documentation style, follow-up length, and mortality density; CME is the strongest correlate of AUPRC we can identify within this design, but it cannot be isolated from the co-varying factors. Compute-matched isoFLOP ablations that independently vary CME, token count, and patient count at fixed compute are needed to establish the causal contribution of outcome resolution, and are the focus of follow-on work.

Two further limits bound the generality of the CME finding. First, CME is defined for mortality and may not transfer directly to non-fatal or episodic endpoints, which may require alternative notions of “resolved” trajectories (for example, episode-years for heart failure or depression). Second, all results are at a fixed model size (1.62 billion parameters); larger architectures may alter the relative importance of data composition versus scale. Nonetheless, CME can be estimated from any EHR system with linked mortality records, giving health systems a practical way to assess whether their longitudinal data are likely to support this approach before investing in pre-training.

### Limitations and future directions

Several limits bound the generality of these findings. First, all documentation comes from a single integrated system with its own templates, coding practices, and predominantly older, male population. VA-LLM is therefore best viewed as a within-system foundation model, and external validation on non-VA populations is essential before broader deployment.

Second, clinical text may encode historical inequities in access, documentation quality, and treatment. Prior work on CAN has identified racial disparities and performance drift under distribution shift^32,33^, and a text-based model trained on the same population may inherit or amplify such biases. Subgroup, reclassification, and recalibration analyses are reported in Supplementary Notes S3.1–S3.3; comprehensive stratified evaluation across facilities, regions, and operational deployment contexts is the focus of follow-on work.

Third, we focus on one outcome family: all-cause mortality at multiple horizons. Mortality likely represents an upper-bound test case for label-efficient trajectory modeling: it is clinically salient, well ascertained through NDI linkage, temporally consequential (a patient’s entire record contributes diffuse prognostic signal), and relatively high-prevalence compared with many actionable clinical endpoints. The label-efficiency gains and scaling patterns reported here may not transfer to outcomes that are rarer, episodic, or intervention-sensitive, such as suicide, 30-day readmission, or disease-specific progression, where the relevant textual signal may be sparser and more localized in time. Establishing whether longitudinal text pre-training offers comparable advantages for these harder endpoints is the most important test of the framework’s generality.

Fourth, VA-LLM processes up to eight 2,048-token chunks per patient without cross-chunk attention, aggregating independent segment representations at inference. Strong discrimination from approximately 16,000 recent tokens suggests that much prognostic signal resides in outcome-proximal text; longer-context or hierarchical architectures could better exploit full-trajectory structure, and multimodal extensions combining text with structured diagnoses, laboratory values, and vital signs may further sharpen the representation of patient state.

## Methods

### Ethical Approval

This study was approved by the VA Central Institutional Review Board. All analyses were conducted within VA- and DOE- approved secure computing environments. The study used retrospective, de-identified data and was granted a waiver of informed consent. Data access was governed by a VA data use agreement and DOE interagency agreement for use of OLCF computing resources.

### Data Source

Clinical text was extracted from the VA’s CDW Text Integration Utility (TIU) database spanning January 1, 2000 through December 31, 2021, encompassing 14,062,540 patients with documented notes. TIU records include progress notes, discharge summaries, consult notes, and other documentation generated during patient encounters. The final pre-training corpus comprised 6.31 terabytes of text from 4.19 billion encounters across 13,783,414 patients; the remaining 279,125 patients (2% stratified sample) were reserved for independent evaluation. SQL extraction scripts are provided in Supplementary Materials.

### Outcome Definitions

#### All-Cause Mortality

Mortality outcomes were ascertained by linking VA clinical records with the CDC’s NDI through December 31, 2021, providing date and cause of death for 7.8 million patients. Each patient was assigned a binary outcome label OC ∈ {0, 1}, where OC = 1 denotes NDI-confirmed death, and an outcome date OC_date_ equal to the NDI-recorded date of death for deceased patients. Surviving controls were assigned an outcome date at the same age as their matched case’s death, as described under Prediction Time Point.

For the model comparison experiments, patients in the 279,125-patient held-out cohort were eligible if they had any clinical text before January 1, 2017. Among eligible patients, those with outcome dates before this cutoff formed the retrospective fine-tuning set (*n* = 144,520), and those with outcomes on or after that date formed the prospective evaluation cohort (*n* = 107,555). The 2017 cutoff was chosen to guarantee at least five years of NDI follow-up (through December 2021) for all prospective patients, ensuring that five-year mortality labels are ascertained rather than right-censored. Within the fine-tuning set, case-control samples of 1K, 10K, and 100K patients were drawn by stratified random sampling, maintaining the observed case-to-control ratio (approximately 1:1) at each size. Each case-control pair was used in a single fine-tuning cohort; controls were not shared across sample sizes or horizons. Characteristics are reported in Table 1.

#### CAN Score

The Care Assessment Need (CAN) score is a validated logistic regression model that estimates 90-day and 1-year mortality risk from structured EHR variables including demographics, diagnoses, utilization patterns, laboratory values, and vital signs^30^. Originally validated on 4.6 million patients with an AUC of 0.86 for one-year mortality, CAN was subsequently refined from 90 to 36 input variables while maintaining discriminative performance^4,30^. Scores are recomputed weekly for all Veterans assigned to a primary care team and used operationally across all VA facilities for care prioritization and palliative care referrals^4^. The version used in this study corresponds to the model parameters in production as of the prediction date.

For clinical validation, CAN 90-day mortality probability scores were obtained as of January 1, 2023 for all nationally- indexed Veterans with active enrollment and at least one clinical note during 2022 (*n* = 5,496,915). Because the NDI linkage ends December 31, 2021, 90-day outcomes for this cohort (deaths occurring January through March 2023) were ascertained from CDW vital status records rather than NDI. This cohort has no overlap with the fine-tuning set and falls entirely outside the pre-training corpus (which ends January 2022), providing a temporally out-of-distribution evaluation. VA-LLM predictions were generated for the same patients using the checkpoint 16 model fine-tuned on 100K patients, with input restricted to clinical text from the preceding year, January 2022 through December 2022.

### Tokenization and Preprocessing

PII including Social Security numbers, phone numbers, patient names, and geographic identifiers were systematically replaced with a `<|censored|>` token using rule-based pattern matching validated through manual review. Minimal preprocessing was applied to remove non-informative elements: redundant whitespace was normalized, and sequences of non-alphanumeric characters, typically demarcating section headers, were removed. A domain-specific byte-pair encoding (BPE) tokenizer was trained on the de-identified pre-training corpus using the Hugging Face `tokenizers` library. Two structural tokens were introduced to maintain document organization: `<|visitbreak|>` to delineate distinct encounters and `<|section|>` to preserve structural breaks such as new paragraphs, section headers, and list items. The resulting vocabulary comprised 94,720 unique tokens.

Clinical notes for each patient were chronologically ordered from first encounter through their final recorded visit or confirmed death during the observation period. Notes generated during the same encounter were concatenated and prepended with `<|visitbreak|>` to mark the start of the encounter. The model pre-trains on one document per patient, where document length reflects the full extent of that patient’s longitudinal care. Median document length was 36,900 tokens (mean 125,900), spanning a median of 52 encounters per patient. Document length varied substantially across birth-year cohorts, with per-cohort mean token counts ranging from approximately 30,000 (earliest cohort, WWII-era Veterans) to 185,000 (middle cohorts with the longest follow-up); the distribution is reported in Supplementary Table S1. Because most documents far exceed the 2,048-token context window, pre-training sees each document as a sequence of context-length segments processed with standard causal attention; a median-length document spans approximately 18 such segments, and the model never attends across segment boundaries during pre-training. A natural extension is to scale the context length to accommodate an entire patient record within a single effective batch, enabling full cross-encounter attention; we leave this to future work.

### Model Architecture and Pre-training

#### Pre-training Curriculum

All nationally-indexed VHA patients were sorted by `BirthDateTime` and partitioned at every one million unique patient identifiers (`PatientICN`). Of each million, approximately 600,000 had documented clinical text, yielding 21 sequential cohorts. Tokenized data were prepared per cohort within the KDI enclave and transferred to Frontier for training. We modified the GPT-NeoX training framework to checkpoint after each cohort rather than at fixed step intervals, producing 21 checkpoints that each correspond to a distinct cumulative composition of the pre-training corpus (Supplementary Table S1).

#### Cumulative Experience

For each patient we compute an observed follow-up duration as the time between the first and last clinical document in the pre-training corpus. Cumulative experience at checkpoint *k* is the sum of these durations across all patients included through that checkpoint. We partition cumulative experience into two complementary components. Cumulative mortality experience (CME) sums follow-up durations for patients with an NDI-confirmed death record:

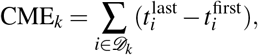

where *D*_*k*_ is the set of patients with NDI-confirmed deaths included through checkpoint *k*, 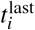 is the date of the last clinical document, and 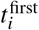 is the date of the first. Its complement, cumulative survival experience (CSE), sums follow-up durations for patients without an NDI death record. CME captures the density of outcome-resolved trajectories in the corpus; CSE captures right-censored follow-up from patients whose vital status is unknown. Both quantities can be estimated from any EHR data warehouse with linked mortality records, without requiring model training or access to clinical text.

#### Architecture and Training

VA-LLM is an autoregressive transformer implemented using the GPT-NeoX architecture^34^ with modifications from Yin et al.’s FORGE models^35^ optimized for Frontier. The model comprises 1.62 billion parameters: 24 transformer layers (*L* = 24), hidden dimension *d* = 2,064, 24 attention heads (*h* = 24, head dimension *d*_*h*_ = 86), and a maximum context length of 2,048 tokens. Rotary positional embeddings encode token position. The 2,048-token context window is short relative to typical patient records (median 36,900 tokens); the sliding-window inference strategy described below recovers coverage of longer histories at the cost of independent chunk encoding. This configuration was chosen as the largest model that permitted routine fine-tuning and inference on a single DGX node (8× A100 80GB) within the VA’s secure KDI enclave.

Pre-training used the LAMB optimizer with a peak learning rate of 0.004 and cosine decay schedule (1% warmup), gradient clipping (maximum norm 1.0), weight decay (0.01), DeepSpeed ZeRO Stage 1 with bfloat16 precision and activation checkpointing on 64 Frontier nodes (512 AMD Instinct MI250X GCDs). Effective batch size was 2 million tokens. Each patient document was seen exactly once (single-epoch training); no data were repeated across or within cohorts.

#### Computational Infrastructure

All raw clinical text remained within the Knowledge Discovery Infrastructure (KDI), a secure VA computing environment hosting two NVIDIA DGX nodes (8× A100 80GB each). Only encoded token sequences, devoid of readable text and patient identifiers, were transferred to Frontier’s CITADEL Secure Protected Infrastructure (SPI)^36^. The VA-specific tokenizer remained isolated within the KDI enclave, preventing decoding on Frontier. Model checkpoints were transferred back to the KDI for fine-tuning and inference.

### Fine-tuning and Evaluation

#### Prediction Time Point

To prevent information leakage from post-outcome documentation, each patient was assigned a prediction time point (*T*_pr_) beyond which no clinical text was used as input. For cases (deceased patients), *T*_pr_ was set to one week before death. Each control was matched 1:1 without replacement to a case on birth date (ties broken randomly); no additional matching constraints (sex, race, facility) were applied. Each control was assigned a *T*_pr_ at the same age as the matched case’s death, ensuring comparable lengths of clinical history. For prospective evaluation, *T*_pr_ was set to January 1, 2017 for the model comparison experiments and January 1, 2023 for the CAN validation (with input restricted to clinical text from the preceding year; see CAN Score under Outcome Definitions).

#### Input Representation

Because patient records typically exceed the 2,048-token context length, a sliding-window approach segmented each record into non-overlapping, contiguous chunks of 2,048 tokens, proceeding backward from *T*_pr_ so that the most recent documentation is always included. We chose eight chunks (approximately 16,000 tokens) as the maximum per patient, balancing coverage of recent clinical history against the computational cost of scoring millions of patients on available hardware. For the median patient, eight chunks cover approximately 44% of the full record. Patients with fewer than eight chunks used all available text. Each chunk is encoded independently with no cross-chunk attention, matching the segment-level processing used during pre-training.

#### Training

A linear classification head (**W**_*c*_ ∈ ℝ^*d×*2^) was applied to each chunk’s final hidden-state representation, and models were trained with binary cross-entropy loss. Low-Rank Adaptation (LoRA)^29^ was applied to the query, key, value, and output projection matrices in all attention layers, keeping base weights frozen. For each model, the LoRA rank *r* was set to yield approximately 100 million trainable parameters (scaling factor *α* = 2*r*), ensuring comparable adaptation capacity across architectures despite differences in hidden dimension and layer count. Hyperparameters were fixed across all experiments: learning rate 1 × 10^*Δ*3^ for 1K-patient cohorts and 1 × 10^*Δ*4^ for 10K and 100K cohorts, effective batch size of 16,384 tokens, AdamW optimizer with weight decay 0.01 and gradient clipping, cosine decay to 10^*Δ*6^ over one epoch, and early stopping if validation loss did not improve by *>* 10^*Δ*4^ over 100 steps. Training used bfloat16 precision.

#### Inference

Each chunk *j* for patient *i* produced logits **z**_*i, j*_ = *f*_*θ*_ (**x**_*i, j*_). Patient-level predictions were computed by averaging logits across chunks and applying softmax:

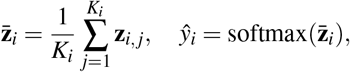

where *K*_*i*_ ≤ 8 is the number of available chunks for patient *i*.

### Comparison Models

Three comparison models were selected to span distinct pre-training domains, all sharing a 2,048-token context window with VA-LLM to permit controlled comparison without architectural confounds: BioGPT_large (1.57B parameters, ∼15M PubMed abstracts), GatorTron_medium (3.91B parameters, *>*90B tokens of University of Florida Health clinical notes), and Llama-2 (7B parameters, ∼2T tokens of general internet text). Pre-trained checkpoints were obtained from Hugging Face^37^: `microsoft/biogpt-large`, `UFNLP/gatortron-medium`, and `meta-llama/Llama-2-7b-hf`. All models were fine-tuned with the identical LoRA configuration, input preprocessing, and hyperparameters described above. Selection rationale and deployment constraints are described in the Results.

### Statistical Analysis

#### Discrimination

Discrimination was evaluated using AUPRC and AUROC. AUPRC served as the primary metric given class imbalance at longer horizons. For VA-LLM, softmax output probabilities were used directly; for CAN, calibrated probability scores were used. Bootstrap 95% confidence intervals (1,000 resamples, percentile method, resampled at the patient level) are reported for all 100K fine-tuning rows in Table 2 and for the CAN comparison in Table 3. Statistical comparison of AUROC between VA-LLM and CAN used the DeLong test^38^. Analyses used scikit-learn.

#### Calibration

Brier scores quantified overall calibration as the mean squared error between predicted probabilities and observed binary outcomes. Calibration slope and intercept were estimated by fitting a logistic regression of the binary outcome *y* on the logit-transformed predicted probability: logit(*P*(*y* = 1)) = *a* + *b* · logit(*p*_pred_), where *b* = 1.0 and *a* = 0.0 indicate perfect calibration. Calibration curves plotted observed mortality rates against mean predicted probabilities within decile bins.

#### Survival Analysis

Patients were stratified into risk quintiles based on VA-LLM predicted probabilities and, separately, CAN scores. For the model comparison experiments, time origin was January 1, 2017 with follow-up through December 31, 2021; for the CAN validation, time origin was January 1, 2023 with follow-up through March 31, 2023 using CDW vital status records. Patients alive at end of follow-up were right-censored. Kaplan-Meier curves were constructed for each quintile, log-rank tests compared survival distributions across strata, and Cox proportional hazards models estimated the hazard ratio for Q5 relative to Q1. The proportional hazards assumption was assessed using Schoenfeld residuals. Survival analyses used `lifelines`.

#### Scaling Variable Analysis

To test whether the clinical composition of the pre-training corpus predicts downstream performance better than corpus size alone, we compared six candidate scaling variables, each computed cumulatively through each of the 21 pre-training checkpoints: total tokens, total patients, total person-years, number of confirmed deaths, CME, and CSE. For each variable *x*, we fit a log-linear regression:

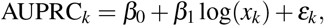

where *k* indexes the 21 checkpoints, and report the coefficient of determination (*R*^2^).

Because all six variables increase together as cohorts are added, a higher *R*^2^ alone does not establish that one variable is a meaningfully better predictor than another; the variables share most of their variation. We therefore applied the Williams test^39^, a method for comparing two correlated predictors that share a common outcome variable measured on the same observations. The test asks whether log(CME) correlates more strongly with AUPRC than log(Tokens) does, accounting for the correlation between the two predictors themselves. We report a one-tailed *p*-value for the alternative hypothesis that CME is a stronger correlate. The full derivation of the test statistic and per-checkpoint variable values are provided in Supplementary Note S3.4.

## Data Availability

Data used for performance analysis in the present study are available upon reasonable request to the authors.

## Competing Interests Statement

The authors have no competing interests to declare.

### Contributorship Statement

R.Z.R. was the software engineer and architect of the VA-LLM and worked on all steps of development and application from pre-training to fine-tuning to inference. J.Y. developed FORGE, which was used as the code base for the development of the VA-LLM on Frontier. S.C. worked on project conception, conceptualization, and supervision. R.Z.R. prepared the paper with reviews and editing by S.C. N.A.K. and J.C.B. worked on supervision.

## Acknowledgments

The authors would like to thank Dr. Sumitra Muralidhar for her invaluable support throughout the development of the VA-LLM. We also thank the MVP Data Core Team and especially Drs. Kelly Cho, Lauren Costa, and Yuk-Lam (Anne) Ho for their support on data safety and transfer. Dr. Ho computed the CAN scores for 5.5 million Veterans. We thank Dr. McMahon and Sayers Dhaubhadel for the many useful discussions on appropriate study designs and statistical validation of the methods. The authors thank the Million Veteran Program (MVP)^40^ staff, researchers, and volunteers who have contributed to MVP, and especially those who previously served their country in the military and now generously agreed to enroll in the study (see `mvp.va.gov` for more information).

