## Supplementary Materials for "A Longitudinal Clinical Foundation Model on Nationwide Veteran Health Trajectories"

[*] A full list of members and their affiliations appears at the end of this document.

#### Contents

- **S1. Supplementary Tables**
  - Table S1: Pre-training curriculum characteristics
  - Table S2: Fine-tuning computational cost
- **S2. Supplementary Figures**
  - Figure S1: Development pathway
  - Figure S2: Corpus properties (outcome completeness and longitudinal coverage)
  - Figure S3: Retrospective–prospective evaluation design
- **S3. Supplementary Notes**
  - S3.1 Recalibration Analysis (Table S3)
  - S3.2 Subgroup Analysis (Table S4)
  - S3.3 Reclassification Analysis (Table S5)
  - S3.4 Scaling Variable Analysis (Tables S6–S7, Figure S4)
- **S4. VA Million Veteran Program Acknowledgements**
- **S5. References**

#### Abbreviations

AUPRC, area under the precision-recall curve; AUROC, area under the receiver operating characteristic curve; CAN, Care Assessment Need score; CDW, VA Corporate Data Warehouse; CME, cumulative mortality experience; CSE, cumulative survival experience (person-years from patients without confirmed deaths); KDI, Knowledge Discovery Infrastructure; LoRA, Low-Rank Adaptation; NDI, National Death Index; TIU, Text Integration Utility; VA-LLM, Veterans Affairs Large Language Model; VHA, Veterans Health Administration.

### S1 Supplementary Tables

Table S1: Pre-training curriculum characteristics across the 21 birth-year-ordered cohorts. **WWII cohorts contribute high mortality with short records; later cohorts contribute longer, more modern records but proportionally more right-censored follow-up.** Patients were sorted by `BirthDateTime` and partitioned at every one million `PatientICN`; approximately 600,000 per cohort had documented clinical text. Early cohorts (c1–c5) contain predominantly WWII-era Veterans with mortality rates exceeding 90%; later cohorts (c16–c21) contain Gulf War and post-9/11 Veterans with mortality rates below 35%. Cumulative TFLOPs estimated as $6ND$ where $N = 1.62 \times 10^9$ parameters and $D$ is cumulative tokens.

| Cohort | Median YoB | Patients | Cum. (M) | Tokens | Cum. (B) | Encounters | Cum. (M) | Person-Years | Cum. (M) | Deaths (cohort %) | Cum. TFLOPS ($\times 10^8$) |
|---|---|---|---|---|---|---|---|---|---|---|---|
| c1:wwii | 1917 | 543,473 | **0.54** | 25.2B | **25.2** | 72.2M | **72.2** | 551K | **0.55** | 541,898 (99.6) | 2.4 |
| c2:wwii | 1921 | 473,711 | **1.02** | 31.7B | **56.9** | 86.8M | **159.0** | 1.97M | **2.52** | 470,046 (99.2) | 5.5 |
| c3:wwii | 1924 | 512,841 | **1.53** | 39.1B | **96.0** | 104.1M | **263.1** | 2.95M | **5.47** | 503,778 (98.2) | 9.3 |
| c4:wwii | 1926 | 564,170 | **2.09** | 49.1B | **145.0** | 126.9M | **390.0** | 3.61M | **9.08** | 543,701 (96.4) | 14.1 |
| c5:wwii | 1928 | 582,811 | **2.68** | 55.7B | **200.7** | 140.6M | **530.6** | 4.34M | **13.42** | 539,537 (92.6) | 19.5 |
| c6:korea | 1931 | 632,629 | **3.31** | 73.2B | **273.9** | 179.6M | **710.2** | 4.85M | **18.27** | 546,585 (86.4) | 26.6 |
| c7:korea | 1934 | 641,239 | **3.95** | 80.1B | **354.0** | 192.0M | **902.2** | 5.83M | **24.10** | 507,895 (79.2) | 34.4 |
| c8:vietnam | 1937 | 636,536 | **4.59** | 74.7B | **428.7** | 181.5M | **1083.7** | 6.21M | **30.31** | 446,301 (70.1) | 41.7 |
| c9:vietnam | 1940 | 655,009 | **5.24** | 95.0B | **523.7** | 225.9M | **1309.5** | 6.32M | **36.63** | 397,749 (60.7) | 50.9 |
| c10:vietnam | 1943 | 681,277 | **5.92** | 113.6B | **637.3** | 264.4M | **1574.0** | 6.57M | **43.21** | 356,380 (52.3) | 61.9 |
| c11:vietnam | 1946 | 665,908 | **6.59** | 118.5B | **755.8** | 226.7M | **1800.7** | 6.42M | **49.63** | 296,029 (44.5) | 73.5 |
| c12:vietnam | 1947 | 735,843 | **7.33** | 139.2B | **895.0** | 321.3M | **2122.0** | 7.32M | **56.95** | 309,430 (42.1) | 87.0 |
| c13:vietnam | 1949 | 703,958 | **8.03** | 127.9B | **1022.9** | 300.1M | **2422.1** | 8.00M | **64.95** | 283,434 (40.3) | 99.4 |
| c14:vietnam | 1951 | 650,446 | **8.68** | 103.2B | **1126.2** | 249.3M | **2671.4** | 7.49M | **72.44** | 265,593 (40.8) | 109.5 |
| c15:vietnam | 1954 | 648,306 | **9.33** | 122.0B | **1248.1** | 290.6M | **2962.0** | 6.61M | **79.06** | 262,469 (40.5) | 121.3 |
| c16:gulf | 1957 | 621,629 | **9.95** | 109.5B | **1357.7** | 262.8M | **3224.8** | 6.72M | **85.77** | 218,046 (35.1) | 132.0 |
| c17:gulf | 1960 | 641,945 | **10.59** | 110.4B | **1468.1** | 258.4M | **3483.2** | 6.29M | **92.06** | 216,617 (33.7) | 142.7 |
| c18:gulf | 1963 | 600,996 | **11.19** | 70.9B | **1539.0** | 166.7M | **3649.9** | 6.38M | **98.45** | 181,155 (30.1) | 149.6 |
| c19:gulf | 1967 | 613,887 | **11.81** | 74.2B | **1613.2** | 169.5M | **3819.3** | 5.44M | **103.89** | 176,096 (28.7) | 156.8 |
| c20:afghanistan | 1971 | 628,254 | **12.43** | 63.7B | **1676.9** | 144.4M | **3963.7** | 5.29M | **109.18** | 180,810 (28.8) | 163.0 |
| c21:afghanistan | 1976 | 643,047 | **13.08** | 45.2B | **1722.2** | 103.4M | **4067.1** | 5.10M | **114.28** | 194,936 (30.3) | 167.4 |

Table S2: Fine-tuning computational cost on a single NVIDIA DGX node (8× A100 80GB) with 100K training patients.

| Model | Pre-training Domain | Parameters | Batch Size | sec/iter | Runtime |
|---|---|---|---|---|---|
| Llama-2 | General | 7B | 8×2048 | 2.57 | 56:38 |
| GatorTron | Clinical (UF Health) | 3.91B | 32×512 | 1.96 | 39:03 |
| BioGPT | Biomedical (PubMed) | 1.57B | 8×2048 | 1.29 | 27:22 |
| VA-LLM | Clinical (VA) | 1.62B | 8×2048 | 0.76 | 15:55 |

#### S2 Supplementary Figures

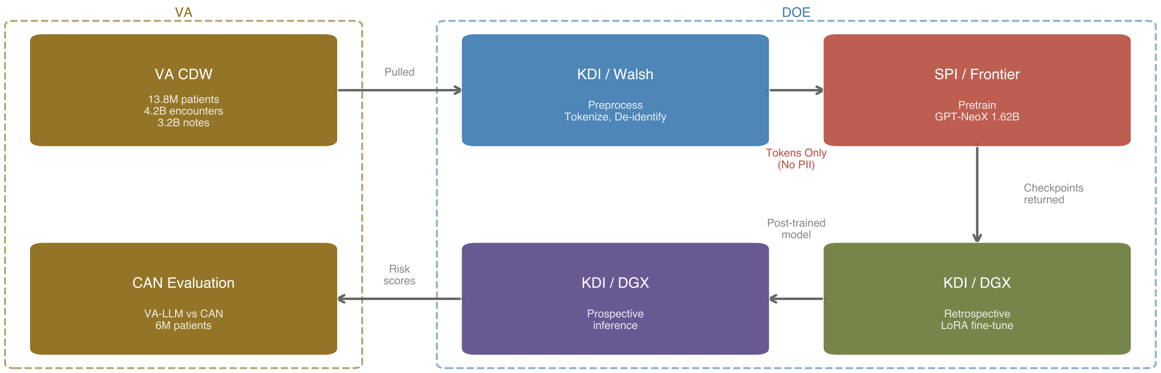


Figure S1: Development pathway. Clinical text was extracted from the VA Corporate Data Warehouse (CDW), transferred to the Knowledge Discovery Infrastructure (KDI) secure enclave, pre-trained on OLCF Frontier, and returned to the KDI for fine-tuning and evaluation.

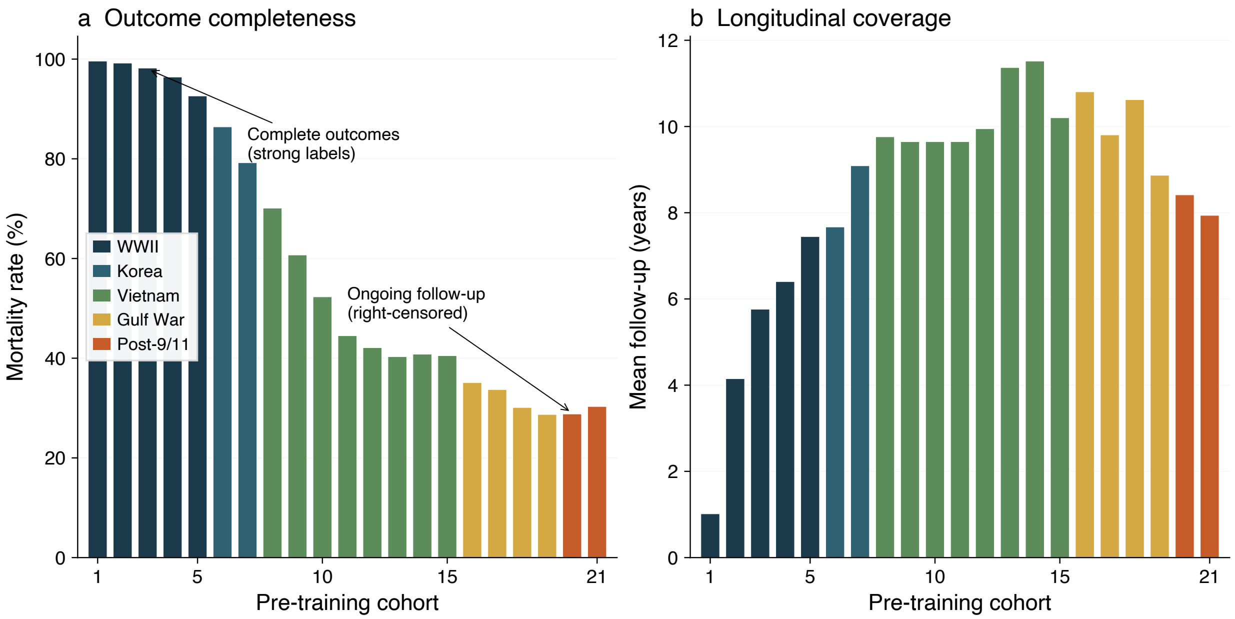


Figure S2: Corpus properties across the 21 birth-year-ordered pre-training cohorts. (a) Mortality rate per cohort: near-complete outcome resolution in early cohorts (99.6% for c1) decreases to approximately 30% for the youngest cohorts, where most patients remain alive and right-censored. (b) Mean follow-up duration per cohort: WWII-era cohorts have the shortest records (high mortality truncates trajectories), while later cohorts have progressively longer observation windows.

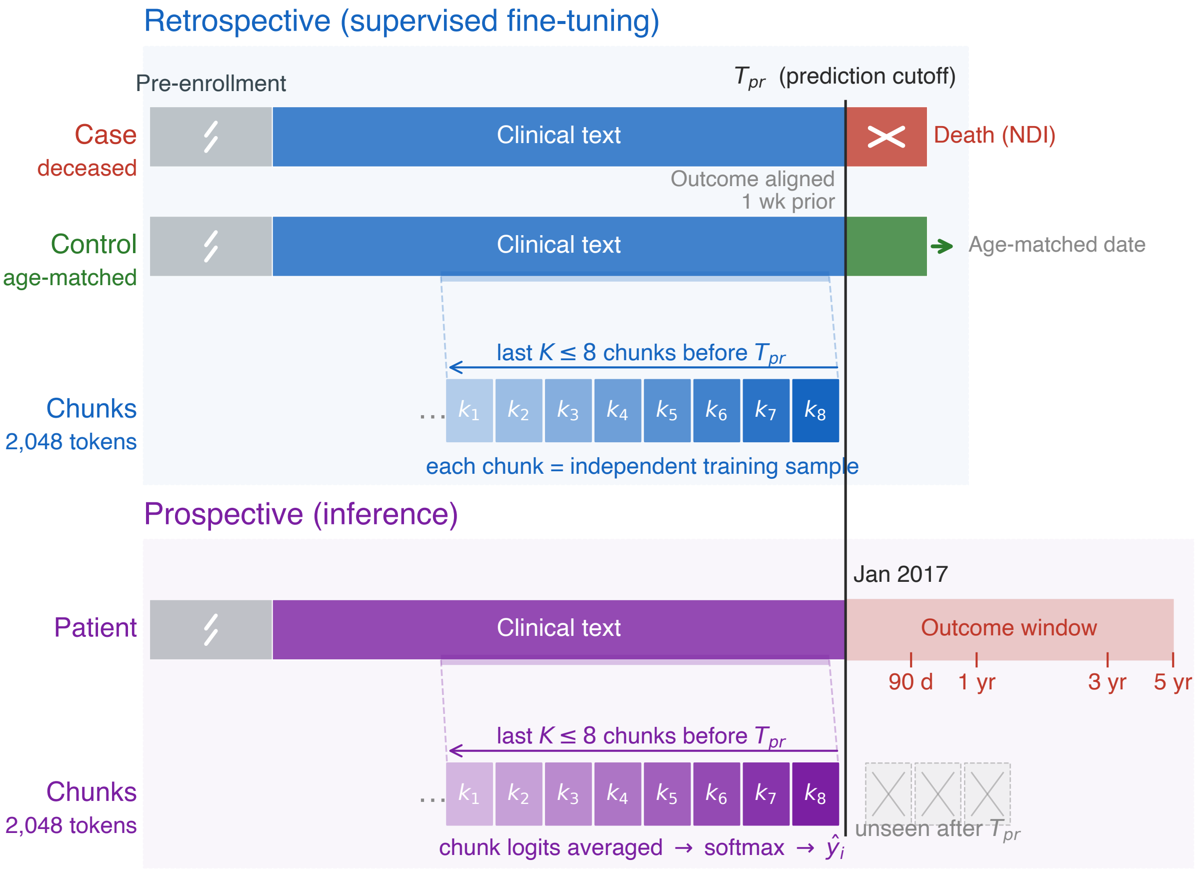


Figure S3: Retrospective–prospective evaluation design. Temporal separation at January 1, 2017 (fine-tuning cutoff) and January 1, 2023 (CAN validation cutoff) ensures that no future information leaks into model training. Top: retrospective case-control fine-tuning with outcome-aligned chunking. Bottom: prospective inference with masked future chunks.

### S3 Supplementary Notes

#### S3.1 Recalibration Analysis

##### Objective

VA-LLM was fine-tuned on balanced case–control samples, producing well-ordered risk rankings but poorly calibrated absolute probabilities (Brier score 0.1554 versus 0.0093 for CAN on the same cohort). This note assesses whether standard post-hoc recalibration recovers absolute-probability accuracy without sacrificing discrimination.

##### Methods

A 5% random subsample of the CAN validation cohort (January 2023) was used to fit an isotonic regression mapping VA-LLM predicted probabilities to observed 90-day mortality rates. The remaining 95% served as a held-out calibration test set on which all post-recalibration metrics are reported. Metrics include Brier score, calibration slope, calibration intercept, AUROC, and AUPRC, before and after recalibration.

##### Results

Isotonic regression fit on 5% of the validation cohort brought the Brier score from 0.1554 to 0.0091, comparable to CAN's 0.0093, while leaving rank-based discrimination essentially unchanged (AUROC 0.8997 versus 0.9000 uncalibrated; AUPRC 0.1450 versus 0.1445 uncalibrated). The calibration slope moved from 0.349 to 0.973 and the intercept from −4.82 to −0.094, both closely matching CAN's calibration (slope 0.995, intercept −0.053). Standard post-hoc recalibration on a small operational sample therefore recovers absolute-probability accuracy potentially suitable for risk communication after local recalibration, while preserving the ranking advantage over CAN reported in the main manuscript (Table S3).

Table S3: VA-LLM calibration and discrimination before and after isotonic regression on the held-out 95% of the CAN validation cohort, with CAN shown for reference. Uncalibrated VA-LLM values match those reported in Table 3 of the main manuscript.

| Metric | VA-LLM (uncalibrated) | VA-LLM (isotonic) | CAN |
|---|---|---|---|
| Brier score | 0.1554 | 0.0091 | 0.0093 |
| Calibration slope | 0.3490 | 0.9725 | 0.9949 |
| Calibration intercept | −4.8232 | −0.0939 | −0.0525 |
| AUROC | 0.9000 | 0.8997 | 0.8774 |
| AUPRC | 0.1445 | 0.1450 | 0.0999 |

#### S3.2 Subgroup Analysis

##### Objective

Aggregate discrimination metrics may mask variation across demographic subgroups. This note reports VA-LLM and CAN performance stratified by sex, race, ethnicity, and age group on the CAN validation cohort (January 2023), with VA-LLM predictions taken from the recalibrated scores of Supplementary Note S3.1. The side-by-side comparison permits assessment of whether text-derived risk estimation generalizes across the subpopulations represented in the VA, and whether either model exhibits stratum-specific deficits relative to the other.

##### Methods

Subgroups were defined as:

- **Sex**: Male, Female.
- **Race**: White, Black, Other/Unknown.
- **Ethnicity**: Hispanic or Latino, Not Hispanic or Latino, Unknown.
- **Age group**: $<55$, 55–64, 65–74, 75–84, $\geq 85$ (at prediction date).

Within each stratum, we computed sample size, observed 90-day mortality, AUROC, AUPRC (both with bootstrap 95% confidence intervals, 1,000 resamples at the patient level), and Brier score, independently for VA-LLM (recalibrated) and CAN.

##### Results

VA-LLM maintained higher AUROC and AUPRC than CAN in every stratum examined, with no reversals or crossings of confidence intervals (Table S4). The AUROC advantage ranged from 1.7 percentage points (Hispanic or Latino) to 5.1 percentage points (age 75–84), and the AUPRC advantage was proportionally larger, ranging from 24% (age $\geq 85$) to 94% (age $<55$) in relative terms. The largest relative AUPRC gains appeared in younger Veterans, where low event rates make precision particularly demanding; VA-LLM nearly doubled CAN's AUPRC in the $<55$ stratum (0.0602 vs. 0.0311). Both models showed declining discrimination with age, consistent with the higher baseline mortality and greater clinical heterogeneity in older cohorts. Across race and ethnicity, the VA-LLM advantage was consistent; we did not observe a stratum-specific performance reversal in these aggregate demographic categories, though finer-grained subgroups were not examined. Brier scores were comparable between VA-LLM and CAN within each stratum, confirming that post-hoc recalibration generalized across subpopulations without requiring stratum-specific adjustment.

Table S4: VA-LLM and CAN 90-day mortality discrimination and calibration by subgroup on the CAN validation cohort (January 2023). VA-LLM values use the recalibrated scores from Supplementary Note S3.1; VA-LLM point estimates reflect checkpoint 16 fine-tuned on 100,000 patients.

| | | | | VA-LLM (recalibrated) | | | CAN | | |
|---|---|---|---|---|---|---|---|---|---|
| Stratum | *N* | Events | Event rate | AUROC, % (95% CI) | AUPRC, % (95% CI) | Brier | AUROC, % (95% CI) | AUPRC, % (95% CI) | Brier |
| *Sex* | | | | | | | | | |
| Male | 4,969,173 | 53,068 | 1.07% | 89.42 [89.31, 89.53] | 13.84 [13.60, 14.10] | 0.0097 | 87.07 [86.94, 87.21] | 10.05 [9.86, 10.26] | 0.0101 |
| Female | 527,061 | 1,432 | 0.27% | 93.77 [93.19, 94.49] | 11.96 [10.51, 13.73] | 0.0025 | 91.61 [90.67, 92.36] | 7.97 [6.78, 9.33] | 0.0026 |
| *Race* | | | | | | | | | |
| White | 3,847,833 | 41,879 | 1.09% | 89.66 [89.53, 89.79] | 13.68 [13.44, 13.94] | 0.0099 | 87.27 [87.12, 87.41] | 9.98 [9.79, 10.20] | 0.0103 |
| Black | 983,999 | 6,453 | 0.66% | 90.16 [89.75, 90.60] | 12.33 [11.60, 13.13] | 0.0061 | 88.13 [87.72, 88.65] | 9.67 [9.12, 10.36] | 0.0062 |
| Other/Unknown | 664,594 | 6,168 | 0.93% | 90.58 [90.21, 91.01] | 13.64 [12.95, 14.32] | 0.0085 | 88.77 [88.44, 89.21] | 10.54 [10.04, 11.31] | 0.0087 |
| *Ethnicity* | | | | | | | | | |
| Hispanic or Latino | 371,065 | 2,179 | 0.59% | 92.60 [92.02, 93.15] | 14.44 [13.13, 15.86] | 0.0053 | 90.86 [90.14, 91.45] | 10.08 [9.18, 11.25] | 0.0055 |
| Not Hispanic or Latino | 4,806,057 | 48,881 | 1.02% | 89.76 [89.64, 89.87] | 13.79 [13.56, 14.05] | 0.0093 | 87.48 [87.33, 87.61] | 9.98 [9.79, 10.17] | 0.0096 |
| Unknown | 319,354 | 3,440 | 1.08% | 89.59 [89.13, 89.97] | 13.57 [12.63, 14.45] | 0.0099 | 87.56 [86.97, 87.97] | 10.26 [9.61, 11.12] | 0.0101 |
| *Age at prediction* | | | | | | | | | |
| $<55$ | 1,472,262 | 1,464 | 0.10% | 85.40 [84.47, 86.32] | 6.02 [5.09, 7.38] | 0.0010 | 81.84 [80.70, 83.16] | 3.11 [2.59, 4.06] | 0.0010 |
| 55–64 | 995,635 | 3,593 | 0.36% | 88.44 [87.87, 88.95] | 9.81 [8.99, 10.63] | 0.0034 | 84.67 [83.95, 85.36] | 6.78 [5.95, 7.48] | 0.0035 |
| 65–74 | 1,446,820 | 12,711 | 0.88% | 86.81 [86.51, 87.14] | 11.97 [11.55, 12.40] | 0.0081 | 82.10 [81.65, 82.48] | 8.16 [7.85, 8.59] | 0.0084 |
| 75–84 | 1,192,670 | 18,054 | 1.51% | 85.02 [84.76, 85.33] | 13.13 [12.76, 13.55] | 0.0138 | 79.92 [79.67, 80.25] | 8.90 [8.60, 9.26] | 0.0144 |
| $\geq 85$ | 388,847 | 18,678 | 4.80% | 75.61 [75.28, 75.99] | 16.10 [15.69, 16.56] | 0.0429 | 72.95 [72.58, 73.24] | 13.00 [12.58, 13.41] | 0.0440 |

#### S3.3 Reclassification Analysis

##### Objective

The main manuscript reports that 187,624 patients were ranked low risk by CAN (Q1–Q2) but high risk by VA-LLM (Q4–Q5), with 523 deaths within 90 days (observed mortality 0.28%); in the converse group, 153,043 patients ranked high by CAN but low by VA-LLM, of whom 205 died (observed mortality 0.13%). This note characterizes these discordant cohorts relative to concordant patients to assess whether the text-based signal that drives reclassification corresponds to clinically recognizable patient phenotypes.

##### Methods

Risk quintiles were computed independently for VA-LLM and CAN on the January 2023 CAN validation cohort. Three comparison groups were defined:

- **Reclassified up** (CAN Q1–Q2, VA-LLM Q4–Q5): patients whom VA-LLM identifies as high risk and CAN does not.
- **Reclassified down** (CAN Q4–Q5, VA-LLM Q1–Q2): patients whom CAN identifies as high risk and VA-LLM does not.
- **Concordant** (same quintile under both models): reference group.

For each group we report patient counts, observed 90-day mortality, and distributions of age, sex, race, and ethnicity (Table S5).

##### Results

The upward-reclassified group was more racially diverse than the downward-reclassified group (21.60% Black vs. 13.05%), an observation that may reflect differences in documentation patterns, disease burden, or coding practices across demographic groups, though targeted follow-up analyses would be needed to distinguish among these explanations. The concordant group (2,582,689 patients) had the highest observed mortality (1.65%), reflecting concentration of true high-risk patients in upper quintiles where both models converge. Age, mortality rate, and clinical interpretation of the discordant groups are discussed in the main text.

Table S5: Characteristics of reclassified and concordant cohorts on the CAN validation cohort (January 2023).

| | Reclassified up (CAN low, VA-LLM high) | Reclassified down (CAN high, VA-LLM low) | Concordant (same quintile) |
|---|---|---|---|
| *N* | 187,624 | 153,043 | 2,582,689 |
| 90-day deaths | 523 | 205 | 42,669 |
| Observed 90-day mortality | 0.28% | 0.13% | 1.65% |
| Median age (years) | 51.74 | 73.87 | 68.36 |
| Sex, % male | 88.94% | 94.30% | 88.98% |
| Race, % White | 64.64% | 76.01% | 70.05% |
| Race, % Black | 21.60% | 13.05% | 17.72% |
| Race, % Other/Unknown | 13.75% | 10.94% | 12.23% |
| Ethnicity, % Hispanic or Latino | 6.88% | 6.55% | 7.04% |
| Ethnicity, % Not Hispanic or Latino | 86.42% | 87.92% | 87.22% |
| Ethnicity, % Unknown | 6.70% | 5.53% | 5.75% |

#### S3.4 Scaling Variable Analysis

##### Objective

The main manuscript reports that cumulative mortality experience (CME) correlates more strongly with 5-year AUPRC than token count or compute. This note provides the formal statistical procedure: the scaling regression across all six candidate variables at each fine-tuning sample size, and the Williams test comparing log-CME with log-Tokens as correlates of downstream discrimination.

##### Methods

For each of the 21 pre-training checkpoints, six candidate scaling variables were computed cumulatively: total tokens, total patients, total person-years, total confirmed deaths, CME (total person-years contributed by patients with NDI-confirmed deaths; see Methods in the main manuscript), and CSE (person-years from patients without confirmed deaths). For each variable $x$, 5-year AUPRC was regressed on $\log(x)$ with ordinary least squares, and the coefficient of determination ($R^2$) and Pearson correlation ($r$) were recorded.

To compare log-CME with log-Tokens as correlates of AUPRC on the same checkpoints, we applied the Williams test (1). Given correlations $r_{ay}$ (log-Tokens with AUPRC), $r_{by}$ (log-CME with AUPRC), and $r_{ab}$ (log-Tokens with log-CME), the test statistic is:

$$t = \frac{(r_{by} - r_{ay})\sqrt{(n-1)(1+r_{ab})}}{\sqrt{2\,\frac{n-1}{n-3}\,|\mathbf{R}| + \left(\frac{r_{ay}+r_{by}}{2}\right)^2 (1-r_{ab})^3}}, \tag{S1}$$

where $|\mathbf{R}| = 1 - r_{ay}^2 - r_{by}^2 - r_{ab}^2 + 2\,r_{ay}\,r_{by}\,r_{ab}$ is the determinant of the $3 \times 3$ correlation matrix and $n$ is the number of observations (checkpoints). Under $H_0 : \rho_{by} = \rho_{ay}$, $t$ follows a $t$-distribution with $n - 3$ degrees of freedom. We report a one-tailed $p$-value for $H_1 : \rho_{by} > \rho_{ay}$. Regressions used `scipy.stats.linregress`; the Williams $p$-values used `scipy.stats.t.cdf`.

##### Results

The CME advantage was consistent at 10K and attenuated at 1K, where only five checkpoints were sampled and all variables were nearly collinear with performance (Tables S6 and S7; Figure S4).

Table S6: Log-linear regression of 5-year AUPRC on candidate scaling variables (Figure 2c in the main manuscript). The full 21-checkpoint series is available at 100K; 10K and 1K span 5 checkpoints (c1, c6, c11, c16, c21). CSE = cumulative survival experience (person-years from patients without confirmed deaths).

| | 100K ($n = 21$) | | | 10K ($n = 5$) | | | 1K ($n = 5$) | | |
|---|---|---|---|---|---|---|---|---|---|
| Variable | $R^2$ | $r$ | $p$ | $R^2$ | $r$ | $p$ | $R^2$ | $r$ | $p$ |
| log(Tokens) | 0.757 | 0.870 | $3.0\times10^{-7}$ | 0.837 | 0.915 | 0.029 | 0.890 | 0.943 | 0.016 |
| log(Patients) | 0.770 | 0.878 | $1.7\times10^{-7}$ | 0.827 | 0.910 | 0.032 | 0.897 | 0.947 | 0.015 |
| log(Person-Years) | 0.852 | 0.923 | $2.5\times10^{-9}$ | 0.885 | 0.941 | 0.017 | 0.895 | 0.946 | 0.015 |
| log(Deaths) | 0.849 | 0.921 | $3.1\times10^{-9}$ | 0.894 | 0.945 | 0.015 | 0.889 | 0.943 | 0.016 |
| log(CME) | **0.906** | **0.952** | $3.4\times10^{-11}$ | **0.921** | **0.959** | 0.010 | 0.880 | 0.938 | 0.018 |
| log(CSE) | 0.812 | 0.901 | $2.5\times10^{-8}$ | 0.884 | 0.940 | 0.017 | 0.878 | 0.937 | 0.019 |

##### Interpretation

The additional variance explained by CME is concentrated in checkpoints where CME and token count diverge most: early cohorts contribute high CME per token (outcome-dense, short documents) while late cohorts contribute low CME per token (longer documents, fewer confirmed deaths). CSE, which captures follow-up from patients without confirmed deaths, explains less variance than CME ($R^2 = 0.812$ vs. 0.906),

Table S7: Williams test: log(CME) vs. log(Tokens) as predictors of 5-year AUPRC. Full 21-checkpoint series, 100K fine-tuning.

| Quantity | Value |
|---|---|
| $r(\log \text{Tokens}, \text{AUPRC})$ | 0.870 |
| $r(\log \text{CME}, \text{AUPRC})$ | 0.952 |
| $r(\log \text{Tokens}, \log \text{CME})$ | 0.973 |
| Williams $t$ | 8.18 |
| $df$ | 18 |
| $p$ (two-tailed) | $< 10^{-6}$ |
| $\Delta R^2$ | 0.149 |

consistent with resolved trajectories being more informative than censored follow-up in this curriculum, though this observational pattern does not establish a causal relationship.

##### Reproducibility

Per-cohort CME is computed as $\text{Deaths}_k \times (\text{Person-Years}_k/\text{Patients}_k)$, cumulated across cohorts. Source data: `data/raw/dataset_summary.csv` (per-cohort curriculum statistics) and `data/raw/finetuned_results.csv` (per-checkpoint AUPRC). Analysis scripts are released with the manuscript code.

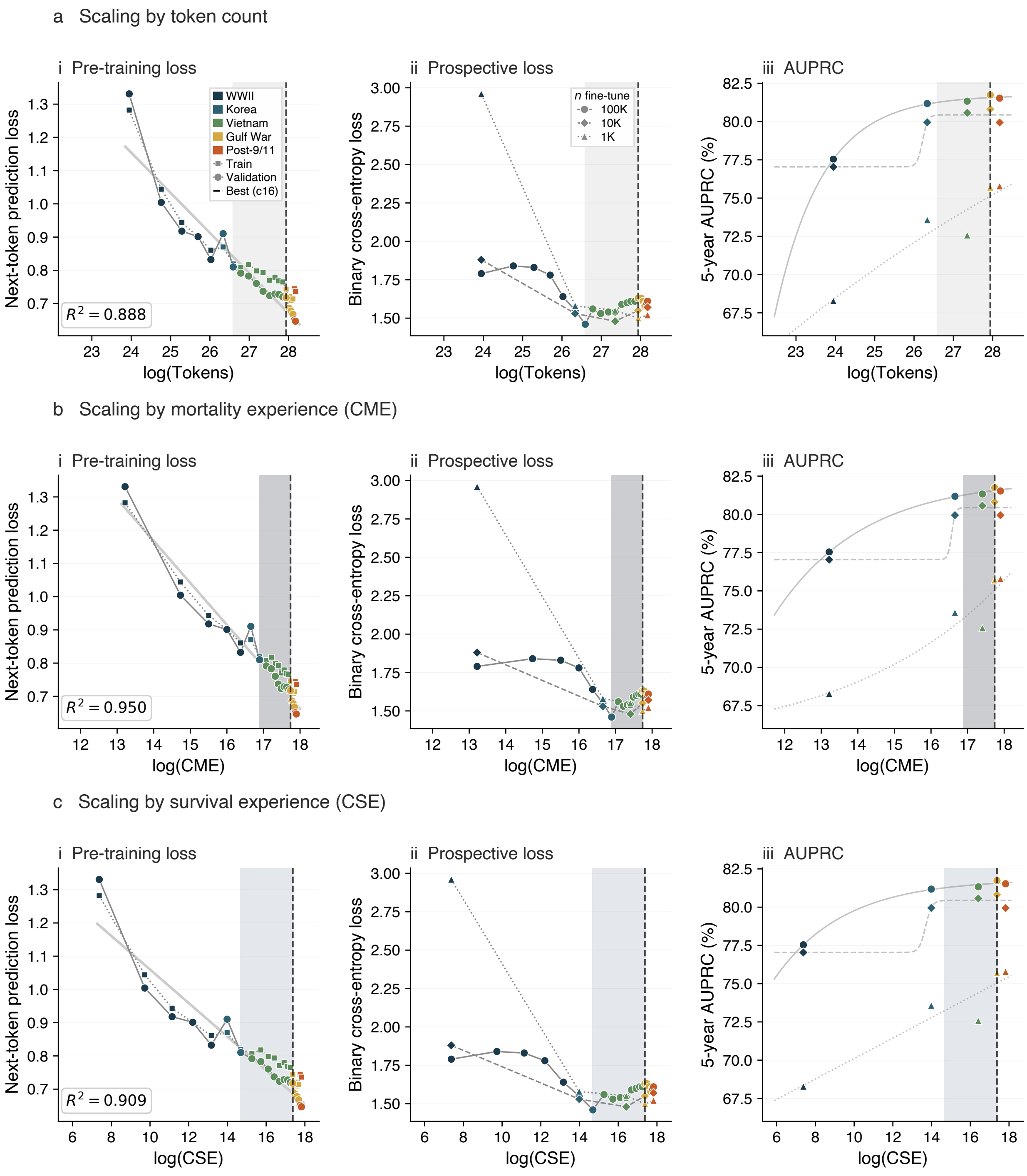


Figure S4: Full scaling grid across 21 pre-training checkpoints. Each row re-indexes the same checkpoints by a different candidate scaling variable: cumulative tokens (a), CME (b), and CSE (c). Columns show next-token prediction loss on the pre-training corpus (i), binary cross-entropy loss on the prospective fine-tuning task at three sample sizes (ii), and five-year AUPRC with logistic S-curve fits at 100,000, 10,000, and 1,000 fine-tuning patients (iii). Pre-training loss decreases continuously across all three axes, but prospective loss and AUPRC plateau after checkpoint 16 (dashed line), paralleling CME saturation. Shaded regions mark the adaptation zone between checkpoint 7 and checkpoint 16; shading intensity scales with $R^2$.

### S4 VA Million Veteran Program (MVP) Acknowledgements


#### MVP Program Office

- Sumitra Muralidhar, Ph.D., Program Director
  US Department of Veterans Affairs, 810 Vermont Avenue NW, Washington, DC 20420
- Jennifer Moser, Ph.D., Associate Director, Scientific Programs
  US Department of Veterans Affairs, 810 Vermont Avenue NW, Washington, DC 20420
- Jennifer E. Deen, B.S., Associate Director, Cohort & Public Relations
  US Department of Veterans Affairs, 810 Vermont Avenue NW, Washington, DC 20420

#### MVP Steering Committee

- Co-Chair: Philip S. Tsao, Ph.D.
  VA Palo Alto Health Care System, 3801 Miranda Avenue, Palo Alto, CA 94304
- Co-Chair: Sumitra Muralidhar, Ph.D.
  US Department of Veterans Affairs, 810 Vermont Avenue NW, Washington, DC 20420
- J. Michael Gaziano, M.D., M.P.H.
  VA Boston Healthcare System, 150 S. Huntington Avenue, Boston, MA 02130
- Adriana Hung, M.D., M.P.H.
  VA Tennessee Valley Healthcare System, 1310 24th Avenue South, Nashville, TN 37212
- Dave Oslin, M.D.
  Philadelphia VA Medical Center, 3900 Woodland Avenue, Philadelphia, PA 19104
- Deepak Voora, M.D.
  Durham VA Medical Center, 508 Fulton Street, Durham, NC 27705

#### MVP Co-Principal Investigators

- J. Michael Gaziano, M.D., M.P.H.
  VA Boston Healthcare System, 150 S. Huntington Avenue, Boston, MA 02130
- Philip S. Tsao, Ph.D.
  VA Palo Alto Health Care System, 3801 Miranda Avenue, Palo Alto, CA 94304

#### MVP Core Operations

- Jessica V. Brewer, M.P.H., Director, MVP Cohort Operations
  VA Boston Healthcare System, 150 S. Huntington Avenue, Boston, MA 02130
- Mary T. Brophy, M.D., M.P.H., Director, VA Central Biorepository
  VA Boston Healthcare System, 150 S. Huntington Avenue, Boston, MA 02130
- Kelly Cho, M.P.H., Ph.D., Director, MVP Phenomics
  VA Boston Healthcare System, 150 S. Huntington Avenue, Boston, MA 02130
- Lori Churby, B.S., Director, MVP Regulatory Affairs
  VA Palo Alto Health Care System, 3801 Miranda Avenue, Palo Alto, CA 94304
- Jacob T. Kean, Ph.D., Acting Director, VA Informatics and Computing Infrastructure (VINCI)
  VA Salt Lake City Health Care System, 500 Foothill Drive, Salt Lake City, UT 84148
- Saiju Pyarajan, Ph.D., Director, Data and Computational Sciences
  VA Boston Healthcare System, 150 S. Huntington Avenue, Boston, MA 02130

- Robert Ringer, Pharm.D., Director, VA Albuquerque Central Biorepository
  New Mexico VA Health Care System, 1501 San Pedro Drive SE, Albuquerque, NM 87108
- Luis E. Selva, Ph.D., Director, MVP Biorepository Coordination
  VA Boston Healthcare System, 150 S. Huntington Avenue, Boston, MA 02130
- Shahpoor (Alex) Shayan, M.S., Director, MVP PRE Informatics
  VA Boston Healthcare System, 150 S. Huntington Avenue, Boston, MA 02130
- Brady Stephens, M.S., Principal Investigator, MVP Information Center
  Canandaigua VA Medical Center, 400 Fort Hill Avenue, Canandaigua, NY 14424
- Stacey B. Whitbourne, Ph.D., Director, MVP Cohort Development and Management
  VA Boston Healthcare System, 150 S. Huntington Avenue, Boston, MA 02130
